# Lifestyle differences between co-twins are associated with decreased similarity in their internal and external exposome profiles

**DOI:** 10.1101/2023.12.12.23299868

**Authors:** Gabin Drouard, Zhiyang Wang, Aino Heikkinen, Maria Foraster, Jordi Julvez, Katja M. Kanninen, Irene van Kamp, Matti Pirinen, Miina Ollikainen, Jaakko Kaprio

## Abstract

Whether differences in lifestyle between co-twins are reflected in differences in their internal or external exposome profiles remains largely underexplored. We therefore investigated whether within-pair differences in lifestyle were associated with within-pair differences in exposome profiles across four domains: the external exposome, proteome, metabolome and epigenetic age acceleration (EAA). For each domain, we assessed the similarity of co-twin profiles using Gaussian similarities in up to 257 young adult same-sex twin pairs (54% monozygotic). We additionally tested whether similarity in one domain translated into greater similarity in another. Results suggest that a lower degree of similarity in co-twins’ exposome profiles was associated with greater differences in their behavior and substance use. The strongest association was identified between excessive drinking behavior and the external exposome. Overall, our study demonstrates how social behavior and especially substance use are connected to the internal and external exposomes, while controlling for familial confounders.

## Introduction

Lifestyle can have a significant impact on health over time, as the years spent in good health increase for individuals who maintain healthy lifestyles (Nyberg et al., 2020). Behavioral risk factors such as smoking, alcohol use and diet account for a substantial part of the global burden of disease (GBD 2019 Risk Factors Collaborators, 2020). These risk factors reflect both societal influences but also genetic predisposition. The same risk factors also influence pathophysiological processes through multiple mechanisms, including metabolic state and gene action.

Omics data enable biological representation at different molecular levels, defining multiple biological layers that can be either deep (e.g., genotype) or shallow (e.g., metabolome). The volume and diversity of omics data increased exponentially since the start of the 21^st^ century (Manzoni et al., 2018) and showed great potential in the study of disease and common traits, as their potential for an in-depth understanding of underlying molecular mechanisms is undeniable (Hasin et al., 2017). As the volume of omics data increases, one concept whose use has recently been on the rise is the *Exposome*, which can be defined as the total environmental exposures that an individual experiences (Wild, 2005). The Exposome is usually classified into three parts: general external, specific external, and internal exposome. The general external exposome generally represents large-scale environmental factors, financial status or the built environment (i.e., green spaces, building density, etc.). The specific external exposome includes specific environmental factors such as diet, occupational exposures and lifestyle factors (Debord et al., 2017). In the context of this paper, we distinguish between the internal and external exposome, the latter including social and physical exposomes (van Kamp et al, 2022) but not lifestyle. Finally, environmental imprints on omic profiles are commonly classified as part of the internal exposome, as omics data has been shown to be highly sensitive to the environment, and genetic factors only explain part of their variance. For example, it has been estimated that 40% to 50% of the variance in metabolite levels is explained by genetics (Pool et al., 2020), and for DNA methylation the mean heritability is 19% with large differences and polygenic effects across methylation sites (van Dongen et al., 2016; Min et al., 2022). This suggests a substantial role of both the genome and environment in one’s omic profile. For example, the epigenome is widely used to estimate various exposures, such as cigarette smoking (Bollepalli et al., 2019; van Dongen et al., 2023), alcohol consumption, as well as one’s biological age (Bell et al., 2019; Duan et al., 2022). While studies focusing on the internal or external exposome are growing in numbers, those aiming at connecting the internal to the external exposome remain scarce (Maitre et al., 2022).

Recently, studies have shown associations between lifestyle and omics data, such as proteomics (Walker et al., 2020; Corlin et al., 2021) and metabolomics (Delgado-Velandia et al., 2022; Kaspy et al., 2022) data. DNA methylation-based biological aging is also being increasingly used as it reflects health-related exposures and cellular processes, thus being more strongly associated with an individual’s health than chronological age. Associations between DNA methylation-based aging with for example diet, obesity, alcohol consumption and physical activity are now well established (Quach et al., 2017; Lundgren et al., 2022; Sillanpää et al., 2019). Recently, Kankaanpää et al. (Kankaanpää et al., 2022) showed that the variation in biological aging that is shared with lifestyle can be explained by both common genetic factors and environmental factors. This finding implies that the molecular mechanisms underlying an individual’s lifestyle are intricate and multi-faceted.

Using and integrating multiple data sources of different types, such as omics or environmental data, could provide valuable insights into the complex relationship that links lifestyle to omics. Multi-omics approaches analyzing multiple omics simultaneously raised the promise of drawing results at the scale of all omic layers, i.e., at a holistic scale, rather than at the scale of within-omic variables (Babu and Snyder, 2023). The use of such approaches, despite the high volume of omics available and their known clinical potential, remains underutilized in the lifestyle literature.

Twin cohorts constitute powerful epidemiological designs, wherein the utilization of twin study designs illuminates the nature of relationships between variables, whether attributable to genetic or environmental factors. As such, twin correlations in monozygotic (MZ) and dizygotic (DZ) pairs are commonly used for such purposes (Boomsma et al., 2002; Posthuma et al., 2003). As MZ co-twins share identical DNA at the sequence level, while DZ co-twins share on average 50% of their segregating genes, higher twin correlations within MZ pairs than within DZ pairs may indicate the presence of genetic effects. Conversely, twin studies enable the detection of environmental factors, as the sum of genetic and environmental effects equal the sum of all measurable and unmeasurable effects (Rijsdijk et al., 2002). A similar approach can be used in bivariate frameworks, using notably cross-twin cross-trait correlations, which may lead to the quantification of genetic or environmental correlations. However, detecting shared genetic and environmental correlations underlying more than a few traits remains challenging. As such, twin correlations are powerful tools to dissect the etiology of traits in univariate or bivariate perspectives, but are not suited for a “whole-dataset” perspective. Consequently, the emergence of multi-omics studies in twin cohorts is challenged by high-dimensional designs. Multi-omics studies in twins are yet expected to improve our understanding of common traits (Hagenbeek et al., 2023), as was shown in few studies of mental health (Zhu et al., 2019), behavior (Hagenbeek et al., 2023), or body weight (Drouard et al., 2023; Bondia-Pons et al., 2014).

To counter the scourge of high dimensions in omics datasets, diverse methods are commonly used in machine learning. As such, multiple clustering algorithms aim to evaluate proximity (i.e., closeness) between individuals at the scale of a whole dataset. Similarity matrices, quantifying between-individual distances, allow to estimate how “close” two individuals are at the scale of a whole dataset (von Luxburg, 2007). Their use, combined with for example Gaussian kernels, can therefore project high-dimensional data into single univariate scores depicting closeness between individuals, called Gaussian similarities. While the use of such an approach is at the root of multiple clustering algorithms, including spectral clustering, we did not find any mention of the use of Gaussian similarities in the literature of twin research.

Our main aim was to investigate whether co-twins differing in lifestyle may also show less similar omic or epigenetic aging profiles, or be exposed to a less similar external exposome, compared to twins from pairs with a very similar lifestyle. To do so, we first quantified, using Gaussian similarities, how close two co-twins are at the scale of a whole dataset (Fig. 1A), across four domains: the plasma proteome, the plasma metabolome, epigenetic age acceleration (EAA), and the external exposome. These scores were later referred to as within-pair proximity scores (WPPS) (Fig. 1B). We then quantified associations between domain-specific WPPS with multiple variables depicting lifestyle, such as education, leisure-time activity, substance use and behavior. We also investigated whether similarity between co-twins at the scale of a domain could be associated with greater similarity at the scale of another domain. Using all twins and subsets of MZ and DZ twin pairs only, we then aimed to investigate whether pairwise WPPS associations were likely to be driven by genetics and/or environment.

**Fig. 1:**
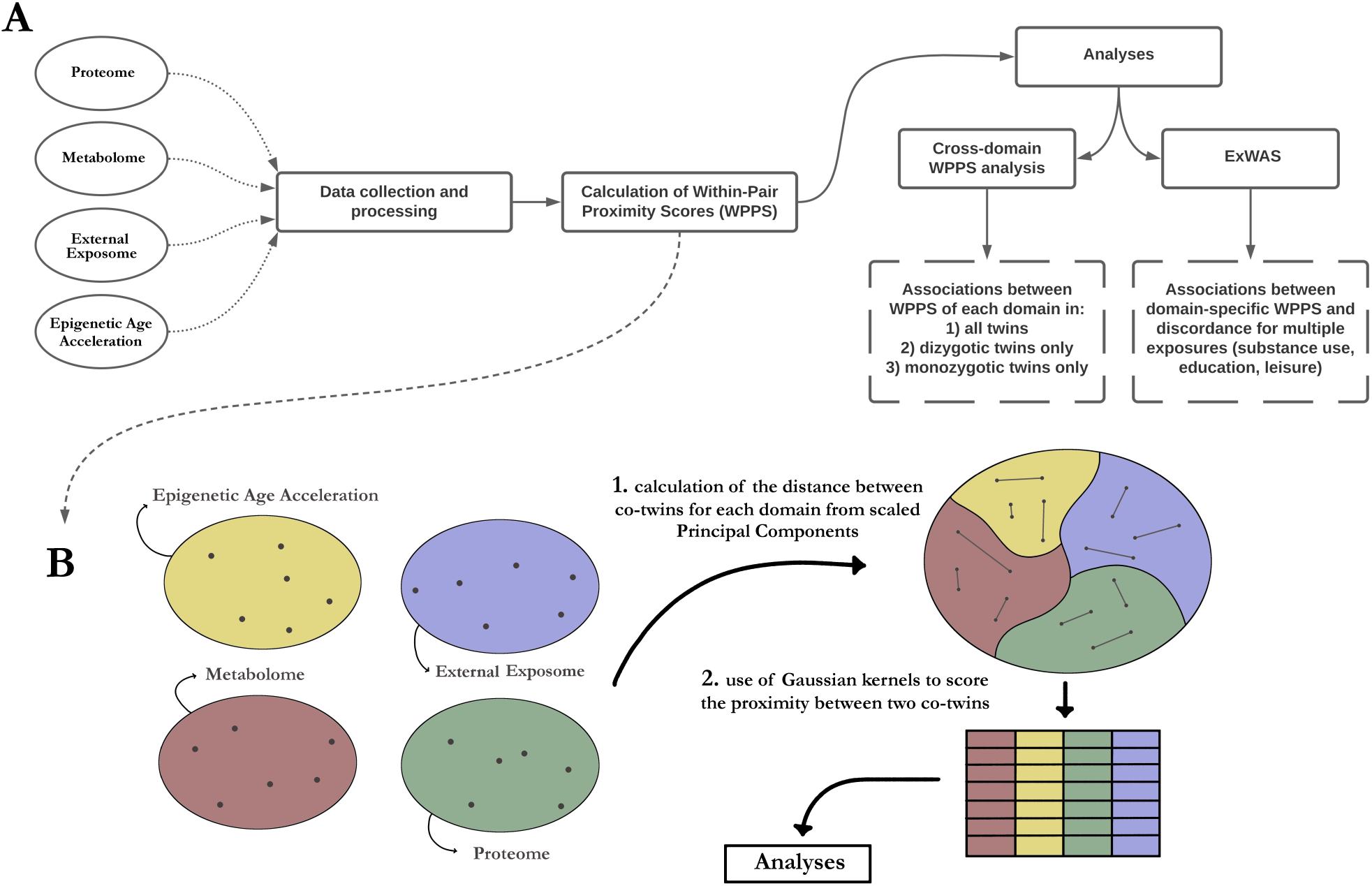
Study workflow diagram. The study was divided into two main analyses **(A)**, which followed the calculation of Within-Pair Proximity Scores (WPPS) at the level of each domain **(B)**. Cross-domain WPPS analysis involved pairwise linear regressions, while the ExWAS analysis was designed to identify associations between lifestyle differences and WPPS scores in twin pairs.

## Results

The present study included up to 257 same-sex twin pairs (63% females; 54% monozygotic pairs) from the FinnTwin12 cohort. The proximity of the twins within each twin pair across the proteome, metabolome, EAA, and external exposome domains was estimated using within-pair proximity scores (WPPS) (Fig. 1; Fig. 2A). MZ pairs had significantly higher domain-specific WPPS for the proteome (p=2.9e-7), the metabolome (p=9.5e-5), and EAA (p=3.5e-8) than DZ pairs, suggesting that genetic factors contribute to co-twin similarity in omic profiles and epigenetic aging. MZ co-twins did not show closer external exposomes than DZ co-twins (p=0.09). Male co-twins had a more similar proteome than females (p-value: 9.8e-4), which was not observed for the metabolome (p-value: 0.18), EAA (p=0.13), and external exposome domains (p=0.70). Age at which the co-twins separated from their familial home was significantly positively associated with greater similarity between the co-twins at the external exposome level (p=4.6e-7) but did not show significant association with domain-specific WPPS for the proteome (p=0.50), the metabolome (p=0.11), or EAA (p=0.88).

**Fig. 2:**
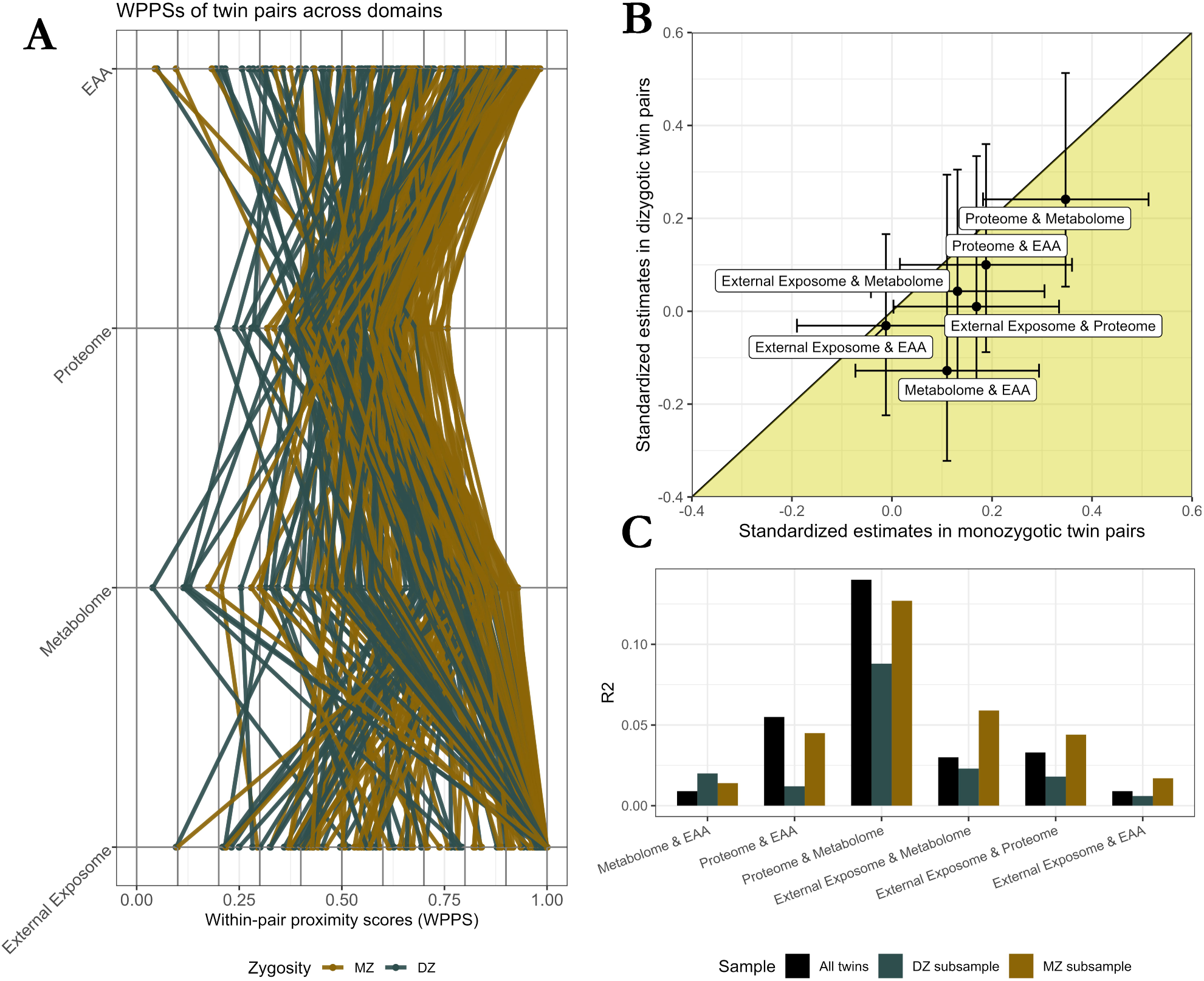
Associations between within-pair proximity scores across the four domains in all twin pairs, in monozygotic twin pairs only, and in dizygotic twin pairs only. **(A)** Graphical representation of the WPPS of each pair across all domains. **(B)** Standardized estimates of the associations across domains for MZ and DZ twin pairs separately. The horizontal and vertical bars represent the 95% confidence intervals of the standardized estimates in the MZ and DZ twin pairs, respectively. Standardized positive coefficients for which the confidence interval does not include zero indicate that a high degree of closeness between co-twins at the level of a given domain was also reflected in a higher degree of closeness between co-twins at the level of a second domain. The range where estimates for MZ twins were larger than those for DZ twins is indicated by the yellow area. **(C)** R^2^ measures in all twin pairs, in MZ twin pairs only, or in DZ twin pairs only. The sample sizes for each of the associations that led to the displayed R^2^ are available in the supplementary material (Figure S1).

### Associations between within-pair proximity scores across different domains

In all twins, greater similarity between co-twins at the level of the external exposome was associated with greater similarity at the level of the proteome (p = 0.04). No significant association was observed between the external exposome with the EAA (p = 0.77) and the metabolome (p = 0.11). The closer the proteomes of two co-twins were to each other, the more similar their epigenetic age acceleration was (p=3.1e-4), while no associations were observed between EAA and metabolome (p=0.25). Finally, greater metabolomic similarity between co-twins was associated with greater proteomic proximity (p=2.6e-8). While most of the significant associations reported are modest in strength (Fig. 2C), about one seventh of the proteome-specific WPPS variance was explained by variations of the metabolome-specific WPPS (R^2^=14.1%). This suggests a substantial interplay between these two omics.

In order to determine whether the associations observed in all twins were likely to be due to genetic or environmental factors, we divided the twin pairs by zygosity into MZ and DZ pairs. For both MZ and DZ pairs, we re-quantified the associations and compared the magnitude of the coefficients by standardizing the WPPS of each domain (Fig. 2B). Number of MZ and DZ pairs with complete WPPS for each domain are available in the supplementary material (Supplementary document 1, Fig. S1). Among MZ twin pairs only, the associations between the proteome and the metabolome (p=5.7e-5), the EAA (p=0.03) and the external exposome (p=0.05) remained significant. All of the coefficients of association between domain-specific WPPS were higher among the MZ twin pairs than among the DZ twin pairs. Similarly, R^2^ measures were larger for MZ than DZ twin pairs for most associations (Fig. 2C). Although this may suggest that genetic factors play a role in within-pair domain associations, we were unable to demonstrate that any of these associations involved genetic factors, as assessed by z-test. The association between the external exposome and the proteome showed the closest level of statistical significance without reaching it. This association remained significant among MZ twin pairs (p=0.05), but not in DZ twin pairs (p=0.92), although the difference in the effects between the two subsamples was not significant (one-tailed z-test: p=0.10).

### Associations between within-pair proximity scores and difference in lifestyle between co-twins

We sought to examine whether lifestyle differences between co-twins were associated with differences between co-twins at the level of each domain. To do so, we quantified associations between WPPS of each domain with 14 lifestyle variables (Fig. 3) that reflected twin discordance (i.e., one co-twin exhibits a feature that the other co-twin does not). The number of discordant twin pairs is available in supplementary material (Supplementary document 1, Table S1).

**Fig. 3:**
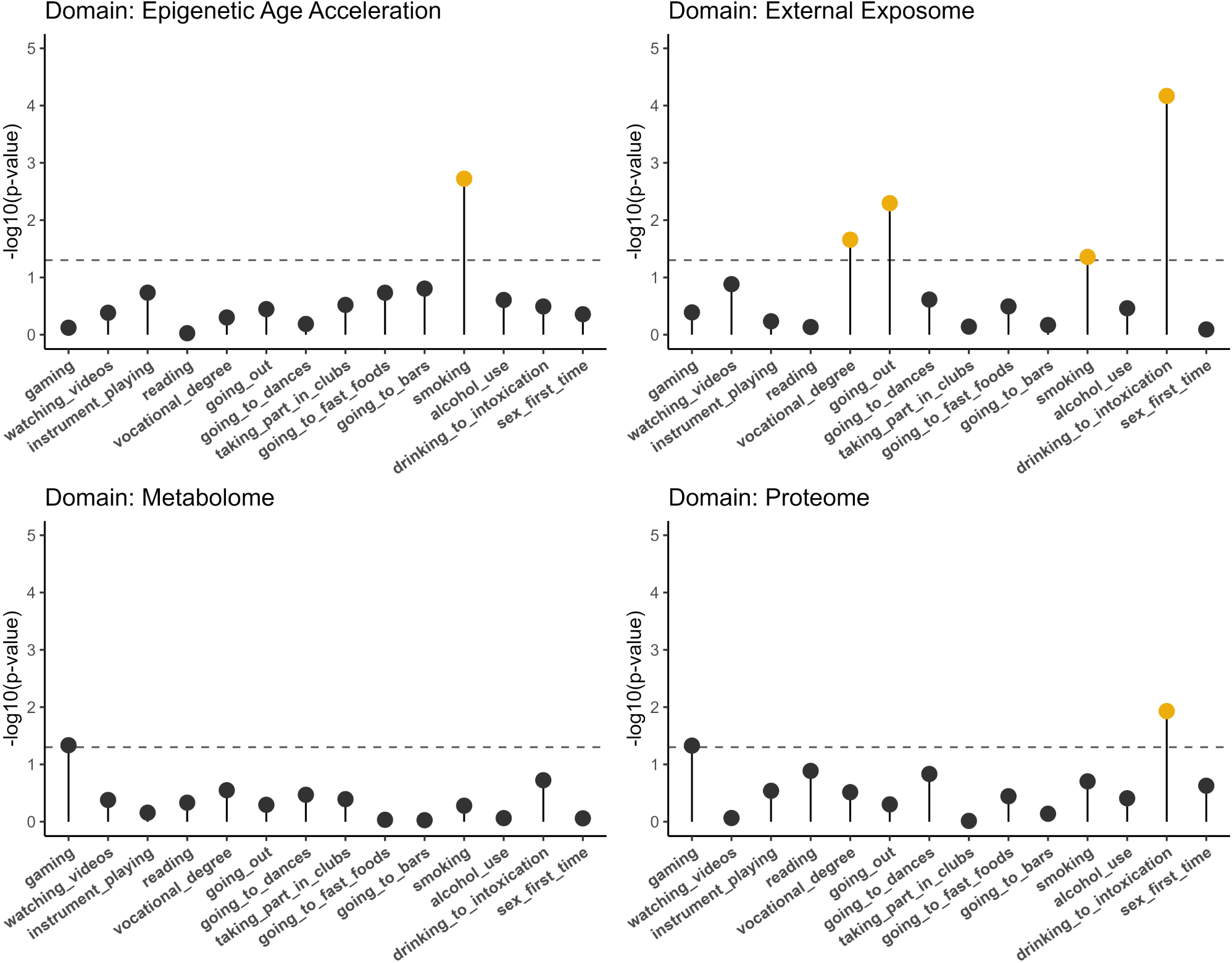
Associations between domain-specific within-pair proximity scores and binary variables indicating discordance in twin pairs. The log_10_ nominal p-values that have FDR corrected p-values of less than 0.20 are shown in yellow. The dashed lines indicate the threshold value of a nominal p-value of 0.05.

Similarity between two co-twins at the metabolomic level, defined by high metabolome-specific WPPS, was not significantly associated with any lifestyle variable. Twin pairs in which one co-twin frequently drinks to the point of intoxication, while the other does not, had a less similar proteome (estimate: -0.04; se: 0.02; nominal p-value: 0.01; FDR-corrected p-value: 0.16) compared to those pairs with twins possessing similar drinking habits.

The more the co-twins differed in their EAA, the greater the chance that the corresponding twin pair was discordant for cigarette smoking (estimate: -0.10; se: 0.03; nominal p-value: 1.9e-3; FDR-corrected p-value: 0.03).

The external exposome captured the most associations with lifestyle variables (Fig. 3). The more the external exposome differed between twins in a pair, the more likely the pair was discordant for lifestyle, such as having a vocational degree(estimate: -0.10; se: 0.04; nominal value: 0.02; FDR-corrected p-value: 0.10), going out (i.e., hanging out)(estimate: -0.10; se: 0.03; nominal p-value: 5.1e-3; FDR-corrected p-value: 0.03), cigarette smoking (estimate: -0.08; se: 0.04; nominal p-value: 0.04; FDR-corrected p-value: 0.15), or frequent drinking to intoxication (estimate: -0.15; se: 0.04; nominal p-value: 6.8e-5; FDR-corrected p-value: 9.5e-4). These associations suggest that both social habits related to substance use and education are linked to the environment in which the twins live. All summary statistics are available as supplementary documents (Supplementary document 1, Table S2).

### Sensitivity analyses

We assessed the reliability of the associations that included the external exposome because of the skewness of its WPPS. Sensitivity analyses were performed by reducing the skewness by 1) transforming the WPPS of this domain or 2) by excluding twin pairs with a WPPS of 1, i.e. pair of twins living in the same household and therefore having the same external exposome. These sensitivity analyses indicate a relatively good reliability of the results of the main analyses, which can be consulted in the supplementary material (Supplementary document 1, Section 1).

## Discussion

By assessing the similarity between twins in a pair in four different domains, we showed that co-twins who differ in lifestyle tend to be less similar at the internal and external exposome levels, despite sharing at least half of their genetic makeup as well as a common familial environment. Specifically, differences in multi-omic profiles within twin pairs were associated with greater within-pair differences in social behavior and substance use. Greatest within-pair differences in the external exposome were identified between co-twins differing in excessive drinking behavior (i.e., frequent drinking to intoxication). These findings put in perspective the multiple layers, from internal to external exposome, that characterize lifestyle differences within twin pairs. In addition, we observed that co-twins with similar external exposomes tended to have more similar proteomes. Greater similarity between co-twins in epigenetic age acceleration or metabolome was also associated with increased similarity in proteome profiles. These findings suggest an interplay in co-twins’ multi-omic resemblance.

Differences in the external exposome between co-twins were strongly associated with differences in lifestyle between the co-twins. Fewer associations were found between lifestyle with the metabolome, the proteome or EAA. This suggests that there is a relatively strong connection among parts of the external exposome. In particular, the results showed that co-twins with a different external exposome tended to differ in terms of social behavior and substance use. This is in line with previous studies showing associations between substance use and the external exposome (Stahler et al., 2013; Galea et al., 2005), as well as between neighborhood organization and alcohol or drug use (Winstanley et al., 2008). Similarly, Williams and Latkin (Williams and Latkin, 2007) found a positive association between neighborhood poverty and substance use. Our study complements these findings in a family setting, where more similar external exposome within twin pairs is associated with more similar behaviors towards excessive alcohol consumption.

Twins share a common family environment as well as half or all of their genetic heritage with their DZ or MZ co-twin, respectively. Thus, by design, a number of measurable and unmeasurable factors shared within each pair are accounted for in the within-pair comparisons. It is of note that the within-pair proximity scores of the external exposome were similar in MZ and DZ pairs, providing empirical evidence to support one central assumption of classical twin modeling, namely that the environmental exposures and experiences are the same, on average, in MZ and DZ pairs. Observed associations are therefore likely to be due to environmental factors specific to each twin, regardless of their zygosity. Similarly, within-family Genome-Wide Association Studies (GWAS) using measured genes can also control for effects such as population stratification and familial biases (Brumpton et al, 2020) compared to our control of genetic effects using twin pairs alone. Interestingly, within-family GWAS analyses revealed a higher SNP heritability for alcohol use than population-based analyses (Howe et al, 2022), while the effect was in the opposite direction for education and smoking in the same analysis. Finally, findings from the current twin study can, to some extent, be extrapolated to a more global family context. DZ twins are genetically matched like normal siblings, although DZ twins may share a more intense early life environment with their co-twins.

Although we provide additional evidence for a direct effect between the external exposome with social behavior and frequent drinking to intoxication, our design does not allow us to establish any causal relationships. High consumption of alcohol is associated with multiple social problems and medical disorders, both mental and somatic. The educational attainment of the young adult twins may for example explain some of the associations between multi-omic profile and lifestyle differences. Additionally, one twin moving away from family home to attend tertiary education may explain within-pair differences in the external exposome, with the latter being a mediator rather than a cause for differences in lifestyles between the co-twins. More studies are needed to disentangle the role of education and socio-economic factors in linking lifestyle and the external exposome.

Metabolites are known to reflect environmental effects (Cai et al., 2020), yet we observed no association between metabolomic and lifestyle similarity. Several factors and limitations of our study may explain the absence of an association. First, the sample size was relatively modest (complete pairs of twins with metabolome-specific WPPS: N = 219), which limited the statistical power. This observation also applies to the other internal exposome domains such as the proteome. However, datasets that include both internal and external exposome data in more than half a thousand twins are rare. Although specific metabolites or classes of metabolites are known to be associated with lifestyle (Kaspy et al., 2022), it remains to be explored whether noticeable associations between the entire metabolome and lifestyle exist. In addition, the set of lifestyle variables used in the current study was relatively limited. For example, we did not include information on physical activity or diet, even though strong connections with some metabolites have been reported in the literature (Kelly et al., 2020; Guasch-Ferré et al., 2018). Whether similarity in co-twins’ metabolome profiles is associated with lifestyle remains inconclusive by the current study.

The similarity of co-twins’ multi-omic profiles was assessed using Gaussian similarities based on modified Euclidean distances. The modification of the Euclidean distance aimed to counteract high data correlations, while adhering to the most classical version of Gaussian kernels. However, other methods could be used to assess the proximity between individuals in a similar context; the Mahalanobis distance is one such method. Using WPPS in contexts with low-dimensional datasets does not seem to be the most effective either. The methodology presented in the current study, although enabling quantification of how similar co-twins are, does not offer an in-depth epidemiological perspective, as genetic modeling does, for example, to estimate variance components. However, the methodology proved effective and useful for within-pair analyses, especially for high-dimensional data. As such, manipulating a univariate score depicting the proximity between twins in a pair is relatively straightforward in simple configurations such as that offered by regression.

In summary, we show here that co-twins with different lifestyles are less similar in their internal or external exposome profiles than are co-twins with similar lifestyles. Results indicate associations between lower similarity in co-twins’ multi-omic profiles and differences in social behavior or substance use, with the strongest association in pairs differing in excessive drinking behavior.

## Methods

### FinnTwin12 cohort and participants

FinnTwin12 is a nationwide cohort based on Finnish twins born 1983-1987, which aims at investigating behavioral development from childhood to adulthood (Kaprio, 2006; Rose et al., 2019). Participants were identified from the population database of the Digital and Population Data Services Agency of Finland (dvv.fi). They completed multiple questionnaires at ages 11/12, 14, 17 and 22. A subset of these twins was studied more intensively, and 786 of them participated in in-person assessments and provided venous blood samples after overnight fasting as young adults (mean: 22.3; sd: 0.6; range: 21-25), from which proteomic, metabolomic and epigenetic data were generated. Besides omics data, we used exposures derived from geocodes to depict each twins’ external exposome (Wang et al., 2023), including for example access to green spaces. Geocodes were based on the full residential history of the twins until the end of 2020, obtained from the Digital and Population Data Services Agency. The metabolome (number of variables: p=140), proteome (p=439), EAA (p=8) and external exposome (p=65) datasets are each referred to as domains from which within-pair proximity scores were later calculated. In addition to domains, we used questionnaire data characterizing education, leisure activities, substance use and behavior. Details about domains and questionnaire data are introduced in eponymous data processing subsections.

### Data processing

The sample sizes of omics and external exposome data varied but overlapped well with most twin pairs having all domains available. The maximum number of available twin pairs was 257 (63% females), all of which had proteomic data and a high proportion of which had data from other domains as well. Details of omics and external exposome overlap are provided in the Supplementary material (Supplementary document 1, Fig. S1).

#### Proteomics

Proteins from the plasma samples of 786 participants were precipitated and subjected to in-solution digestion according to the standard protocol of the Turku Proteomics Facility (Turku Proteomics Facility, Turku, Finland). Details about high abundant protein depletion, precipitation and digestion in this sample have been described elsewhere (Afonin et al., 2023). Samples were first analyzed by independent data acquisition LC-MS/MS using a Q Exactive HF mass spectrometer and further analyzed using Spectronaut software. Data was locally normalized (Callister et al., 2006), and raw matrix counts were processed and quality controlled as described elsewhere (Drouard et al., 2023). Briefly, protein levels were log_2_-transformed and proteins with >10% missing values were excluded. Missing values were imputed by the lowest observed value for each protein carrying missing values. Corrections for batch effects were performed with Combat (Leek et al., 2012) and the final proteomic dataset comprised 439 proteins which were scaled, such that one unit corresponded to one standard deviation (sd) with zero mean. Proteomic data were available for 257 complete pairs of same-sex twins. The list of proteins is presented in the supplementary material (Supplementary document 1, Table S3).

#### Metabolomics

Metabolites were quantified from plasma samples using high-throughput proton nuclear magnetic resonance spectroscopy (^1^H-NMR) (Nightingale Health Ltd, Helsinki, Finland) (Soininen et al., 2015; Bogl et al., 2016; Rose et al., 2019). An extensive description of the data is available elsewhere (Whipp et al., 2022). The data initially comprised 149 metabolites including lipids and lipoprotein subclasses within 14 subclasses, fatty acid composition, and various low-molecular weight metabolites, including amino acids. We excluded pregnant women (n=53) and an individual with cholesterol medication. Only metabolites with less than 10% missing values were kept, leading to a selection of 140 metabolites (see Table S4 for details of the metabolites used)(Supplementary document 1, Table S4). Imputation by the observed sample minimum value was performed for each metabolite (rate of missing values in dataset: 1.2%). None of the participants had at least one of the first three principal components derived from principal component analyses above or below 5 SD of the mean, indicating the absence of outliers. Metabolite values were normalized using inverse normal rank transformation, and scaled so that one unit corresponded to a change of one sd, with a mean of zero.

#### Epigenetic age acceleration

DNA methylation levels were quantified using Infinium Illumina HumanMethylation450K, and preprocessed using R-package *meffil* (Min et al., 2018) removing bad quality samples and probes (Sehovic et al., 2023). We calculated different EAA estimates, defined as the residuals of chronological age regressed on epigenetic age, using their respective algorithms: Horvath (Horvath, 2013), Hannum (Hannum et al., 2013), PhenoAge (Levine et al., 2018), and GrimAge (Lu et al., 2019), alongside their respective PC-score versions (Higgins-Chen et al., 2022). Details about EAA calculations are described elsewhere (Kankaanpää et al., 2022). In contrast with chronological age, which is defined by the amount of time elapsed since birth, epigenetic age aims at estimating cells’ biological age and shows promising clinical potential (Bell et al., 2019). The final EAA dataset consisted of 8 different EAAs available for 241 complete same-sex twin pairs, showing relatively moderate correlations (mean Pearson correlation: r=0.49) between the EAA estimates.

#### External exposome

The domain of the external exposome comprised a total of 65 exposures derived from two sources: Equal-life project (van Kamp et al., 2022) enrichment datasets and Statistics Finland. We used twin’s geocodes in 2005-2006 to merge the exposures, which were extensively described elsewhere (Wang et al., 2023). Briefly, exposures were continuous and included: sizes of and access to green spaces, percentage of built up areas, population ages and headcounts, crime rates, and voting patterns at municipal elections. Complete dataset was available for 250 pairs of same-sex twins. Complete description of the external exposome’s variables is available in the supplementary material (Supplementary document 1, Table S5).

#### Lifestyle variables

A panel of 14 variables characterizing education, leisure activities, substance use, and social behavior, were derived from home questionnaires completed by the twins prior to the in-person study as well as questionnaires completed during the study visit. These variables were used to examine whether these were associated with domain-specific WPPS. Playing video games, watching videos, playing an instrument, and reading were used as variables to characterize leisure time. Social behavior was assessed by the following variables: going out (i.e., hanging out), going dancing, taking part in a club, going to fast food, and going to bars. Variables characterizing substance use were cigarette smoking, alcohol consumption, and alcohol-induced intoxication (Sipilä et al., 2016). These frequency variables were dichotomized into binary variables, such that modalities were defined by a frequency of at most once a month *vs.* at least once a week. In addition, the attainment of a vocational degree (modalities: yes/no) was used to characterize possible educational differences between the co-twins. We also included a variable indicating the age of the first sexual intercourse (modalities: strictly before the age of 18/ at age 18 or after).

Within-pair lifestyle variables were then created. They were coded as follows: 1 indicates a discordant pair, 0 a concordant pair. That is, for each within-pair indicator variable, 1 denoted a twin pair in which one twin expressed a feature that the other did not, and 0 denoted two co-twins expressing the same feature (e.g., same substance use or social behavior). The number of lifestyle discordant twin pairs ranged from 42 to 94 among the 257 available twin pairs. For each variable, pairs of twins in which at least one of the co-twins had a missing value were coded as having a missing value for the pair. Detailed numbers of discordant and missing pairs per each lifestyle variable are available in the supplementary material (Supplementary document 1, Table S1).

### Statistical analyses

#### Within-pair proximity scores (WPPS) across domains

Gaussian kernels, as defined by (Δ), are commonly used functions to score the proximity between two individuals ***x_i_*** and ***x_j_*** by projecting their mutual distance D(*x_i_*, *x*_j_) (e.g., euclidean distance) into the interval]0,1], with σ being a tunable hyperparameter controlling the width of the neighborhoods.

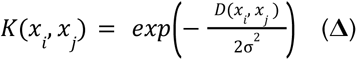

A proximity score of 1 therefore reflects a pair of identical individuals, while a decreasing score value reflects an increased dissimilarity between two individuals. We propose to adapt the Gaussian kernel to our twin design, by scoring the proximity between co-twins, which we refer to as within-pair proximity scores (WPPS). Because correlations between variables within each domain may be substantial, as in the metabolomic data, we did not use the euclidean distance as distance D(.,.), but a weighted version of it. This avoided inflation of proximity scores between two co-twins due to repetition of information from highly correlated variables. For each domain (e.g., metabolome, external exposome, etc.) of dimension *p*, WPPS were calculated as described by the following sequential procedure:

1. A Principal Component Analysis (PCA) was first conducted, from which *p* Principal Components (PCs) were derived, covering 100% of the initial inertia. We denote ω_k_ the percentage of inertia covered by PC_k_ (*k*=1, …, *p*), such that ⅀_k_ω_k_=1.
2. Then, the *p* PCs were scaled to mean zero and variance one.
3. The distance D(*xi*, *xj*) between two co-twins *x_i_* and *x_j_* was defined by weighting each PC by its associated percentage of inertia, i.e., D(*x_i_*, *x_j_*)= ⅀_k_ω_k_[PC_k_(*x_i_*)-PC_k_(*x_j_*)]^2^.
4. The proximity between the two co-twins was calculated as in (Δ). The hyperparameter σ was set to one.

Two co-twins for which the squared differences in levels across all *p* PCs of a domain were higher on average than a standard deviation have a WPPS lower than *exp(-1/2) ≃ 0.61*.

#### WPPS-WPPS and WPPS-lifestyle associations

First, we sought to investigate whether sex, zygosity and age at which the co-twins were separated from the family home at the first time (Wang et al., 2022) were associated with each domain-specific WPPS. We therefore fitted linear regressions modeling a WPPS as dependent variable, and sex, zygosity, and age at separation of the pair as independent variables.

We then examined whether co-twins being similar at the scale of a particular domain may also share greater similarity at the scale of another domain. To do so, we quantified pairwise associations between WPPS across domains using linear regressions. We modeled these associations considering one as the dependent variable, the other as independent variable. The dependent variable for each domain pair was prioritized as follows: 1) external exposome, 2) EAA, 3) proteome, and 4) metabolome. Differences in age at blood sampling between co-twins was added as covariate to correct for potential effects on WPPS, even though these differences were of zero days for three fourths of twin pairs and averaged only to 20 days in all twins. A WPPS of a specific domain was considered significantly associated with a WPPS of another domain if the coefficient of the latter was significantly non-zero, as assessed by t-test (p<0.05).

To assess whether significant associations are likely to be due to genetic effects, we stratified linear regression models by zygosity, as MZ twins are fully matched for genetics, while DZ twins are half-matched. We scaled each WPPS to mean zero and variance one in each subsample, as to be able to compare coefficients across the two subsamples. Nullity of differences in these coefficients between MZ and DZ subsample was assessed by unilateral Z-test (Clogg et al., 1995; Paternoster et al., 2006). Testing was one-sided as MZ twin pairs differ from DZ twin pairs by better genetic matching within the pair; the larger the genetic effects are, the larger the coefficients in the MZ should be compared to those in the DZ twin pairs. Sample sizes for all WPPS-WPPS association scenarios in all twin pairs, MZ pairs only, or DZ pairs only are shown in the Supplementary Material (Supplementary document 1, Fig. S1).

At the second step, we sought to investigate whether co-twins sharing similar proteome, metabolome, EAA or external exposome differed in lifestyle. To do so, we quantified associations between WPPS from each domain with discordance for 14 lifestyle variables, as in a standard Exposome-Wide Association Study (ExWAS). Differences in age at blood sampling were added as a covariate. Associations between each domain-specific WPPS with lifestyle variables were corrected for multiple testing by False Discovery Rate (FDR) using the Benjamini-Hochberg method, and we discussed associations for which FDR-corrected p-values were lower than 0.2.

#### Sensitivity analyses

Because the twins were young, some of them lived in the same familial household as their co-twin. This resulted in 80 twin pairs with an external exposome WPPS of 1. To evaluate potential problems caused by the skewness of this WPPS in linear modeling (skewness: -0.73), we performed two sensitivity analyses. First, we modified the external exposome WPPS by transforming it with the *logit* function so that all the values are more spread out on the whole real axis. As the value 1 is not within the valid range of application of the logit function, we first remapped WPPS into the interval [0.025, 0.975] by linear transformation. The second sensitivity analysis consisted of excluding pairs with a WPPS score of 1 from the analyses in which the external exposome was included. We reported associations in both configurations and discussed potential differences observed with the original modeling.

## Declarations

### Data availability

FinnTwin12 data analyzed in this study is not publicly available due to the restrictions of informed consent. Requests to access these datasets should be directed to the Institute for Molecular Medicine Finland (FIMM) Data Access Committee (DAC) for authorized researchers who have IRB/ethics approval and an institutionally approved study plan. To ensure the protection of privacy and compliance with national data protection legislation, a data use/transfer agreement is needed, the content and specific clauses of which will depend on the nature of the requested data.

### Code availability

Methodological resources, including R scripts, are available upon request from the corresponding author.

### Ethics

The ethics committee of the Department of Public Health of the University of Helsinki and the Institutional Review Board of Indiana University approved the FinnTwin12 study protocol from the start of the cohort. The ethical approval of the ethics committee of the Helsinki University Central Hospital District (HUS) is the most recent and covers the most recent data collection (wave 4) (HUS/2226/2021). The HUS reviews the study annually, and 2023’s statement is number 4/2023, dated 1 February 2023. All participants and their parents/legal guardians gave informed written consent to participate in the study. The authors assert that all procedures contributing to this work comply with the ethical standards of the relevant national and institutional committees on human experimentation and with the Helsinki Declaration of 1975, as revised in 2008.

### Funding

Phenotype and omics data collection in FinnTwin12 cohort has been supported by the Wellcome Trust Sanger Institute, the Broad Institute, ENGAGE – European Network for Genetic and Genomic Epidemiology, FP7-HEALTH-F4-2007, grant agreement number 201413, National Institute of Alcohol Abuse and Alcoholism (grants AA-12502, AA-00145, and AA-09203 to R J Rose and AA15416 and K02AA018755 to D M Dick, and R01AA015416 to J Salvatore), the Academy of Finland (grants 100499, 205585, 118555, 141054, 264146, 308248 to JK, and grants 328685, 307339, 297908 and 251316 to MO, and the Centre of Excellence in Complex Disease Genetics grants 312073, 336823, and 352792 to JK), and Sigrid Juselius Foundation (to MO). This research was partly funded by the European Union’s Horizon 2020 research and innovation program under grant agreement No 874724 (Equal-Life). Equal-Life is part of the European Human Exposome Network.

## Acknowledgements

We gratefully acknowledge the contribution of the Equal-Life consortium at large and the members of the advisory committee.

## Contributions of authors

GD, ZW, and JK conceptualized the study. GD developed the methodology and performed statistical modeling. MF, JJ, KMK, IvK, MO, and JK participated in data acquisition by providing funding and/or technical support. GD, ZW, and AH processed plasma omics and lifestyle data, exposome data, and epigenetic data, respectively. MP, MO and JK supervised the project and/or acquired funding used in this project. GD drafted the first version of the manuscript. All authors contributed to editing the draft, and approved the final version of the manuscript.

## Competing Interests

The authors have no competing interests to declare.

## Notes

### Competing Interest Statement

The authors have declared no competing interest.

