## Supplementary document 1 for "Lifestyle differences between co-twins are associated with decreased similarity in their internal and external exposome profiles"

### Section 1

We investigated the effect of exposome WPPS skewness on the results obtained in analyses involving this domain. Sensitivity analysis 1 consisted in taking the logit of the WPPS of the external exposome to reduce skewness of this variable. Sensitivity analysis 2 consisted in excluding individuals with a WPPS of 1 from the analyses.

**Sensitivity analysis 1:**

The skewness was reduced from -0.73 to -0.31 by using the logit of the WPPS. Pairwise associations between WPPS showed that the association between proteome and external exposome remained positive and significant in all pairs (p=0.04) and MZ pairs (p=0.05), but not DZ pairs (p=0.88). The other associations were not significant. This was consistent with the results of the main analyses.

In the main analyses, we observed associations between the WPPS of the external exposome with vocational degree obtention (estimate: -0.10; se: 0.04; nominal p-value: 0.02; FDR-corrected p-value: 0.10), going out (estimate: -0.10; se: 0.03; nominal p-value: 5.1e-3; FDR-corrected p-value: 0.03), cigarette smoking (estimate: -0.08; se: 0.04; nominal p-value: 0.04; FDR-corrected p-value: 0.15), or frequent drinking to intoxication (estimate: -0.15; se: 0.04; nominal p-value: 6.8e-5; FDR-corrected p-value: 9.5e-4). In the sensitivity analysis, only cigarette smoking was no longer significantly associated with WPPS of the external exposome. However, the FDR-corrected p-value (p=0.21) was close to that obtained in the main analyses (p=0.15). The other three associations remained associated with the external exposome with similar intensity. Frequent drinking to intoxication was the most strongly associated with the external exposome (t-value: -4.2; nominal p= 5.5e-5). Associations between the external exposome with going out (t-value: -3.4; nominal p=7.9e-4) and vocational degree obtention (t-value: -2.7; nominal p=8.4e-4) were found to be even more intense than in the main analyses.

**Sensitivity analysis 2:**

In twin pairs in which the WPPS of the external exposome was not 1 (N pairs=170), the pairwise association between the WPPS of the proteome and that of the external exposome was no longer significant in all pairs (estimate: 0.22; standard error: 0.18; p=0.23) and in MZ pairs (estimate: 0.18; standard error: 0.10; p=0.08). However, the estimates were relatively consistent with those found in the main analyses in all pairs (estimate: 0.31; standard error: 0.15) and in MZ pairs only (estimate: 0.17; standard error: 0.08). These results suggest that a decrease in statistical power due to a reduction in sample size is likely to be responsible for the lack of significance in these associations.

In the main analyses, we observed associations between the WPPS of the external exposome with vocational degree obtention (estimate: -0.10; se: 0.04; nominal p-value: 0.02; FDR-corrected p-value: 0.10), going out (estimate: -0.10; se: 0.03; nominal p-value: 5.1e-3; FDR-corrected p-value: 0.03), cigarette smoking (estimate: -0.08; se: 0.04; nominal p-value: 0.04; FDR-corrected p-value: 0.15), or frequent drinking to intoxication (estimate: -0.15; se: 0.04; nominal p-value: 6.8e-5; FDR-corrected p-value: 9.5e-4). Of these 4 associations, 2 remained significant in twins who did not share the exact same external exposome as their co-twin (i.e., WPPS of 1). As such, frequent drinking to intoxication (t-value: -2.9; nominal p=4.2e-3) and cigarette smoking (t-value: -2.4; nominal p=0.02) were significantly associated with the external exposome. Going out (p=0.10) and vocational degree obtention (p=0.34) were not significantly associated with the external exposome.

### Supplementary Figure

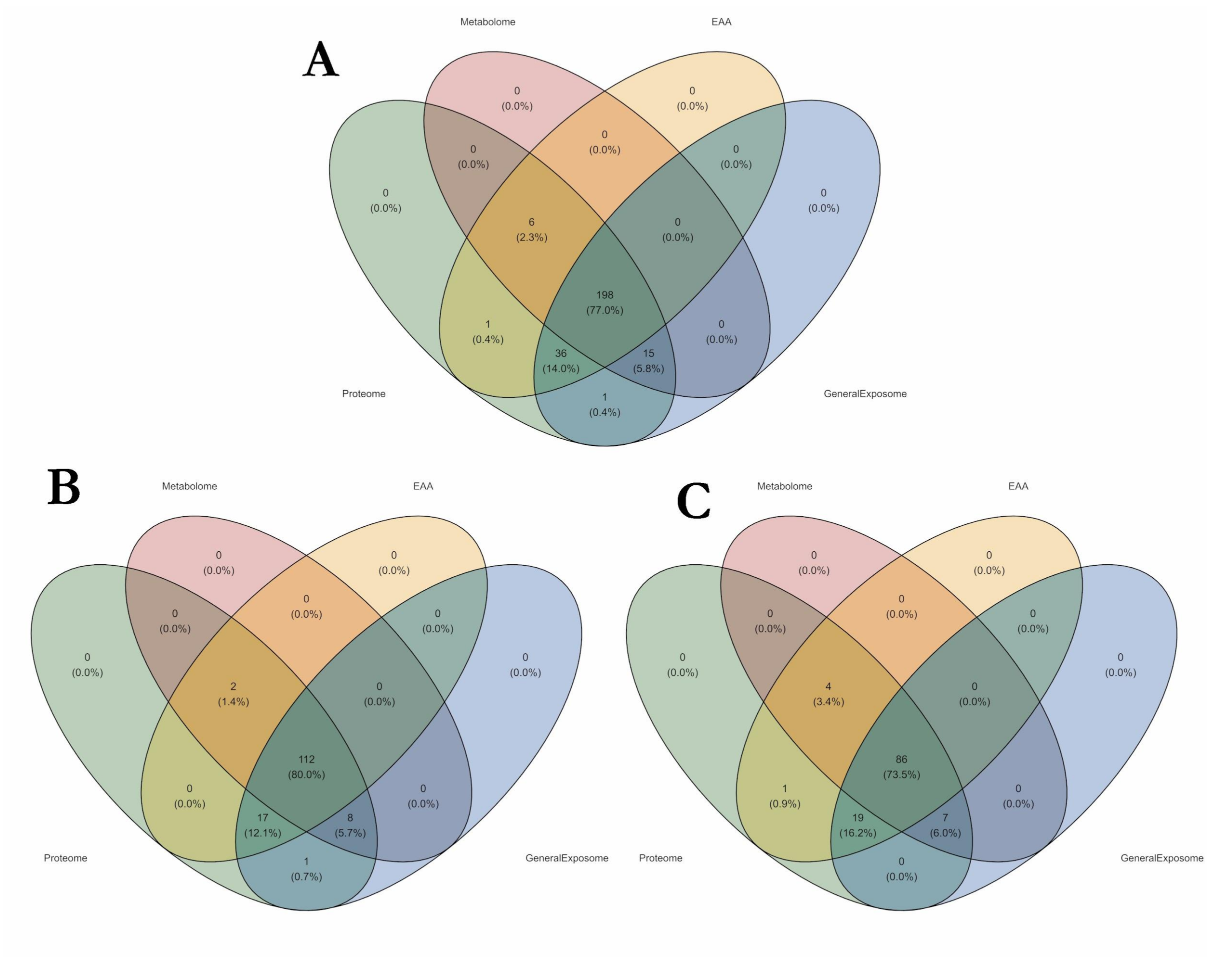


**Fig. S1:** Overlap of samples across domains in all twin pairs, in monozygotic twin pairs only, or in dizygotic twin pairs only.

**Legend: (A)** Overlap of samples in all twins. **(B)** Overlap of samples in monozygotic twins only. **(C)** Overlap of samples in dizygotic twins only.

### Supplementary Tables

**Table S1:** Number of discordant pairs of twins out of the 257 pairs available for the analysis between the domain-specific WPPS and the binary indicators.

| Variable | Number of discordant pairs | Number of NA |
|---|---|---|
| vocational_degree | 46 | 12 |
| watching_videos | 93 | 4 |
| gaming | 76 | 5 |
| instrument_playing | 58 | 5 |
| reading | 94 | 3 |
| going_out | 92 | 4 |
| going_to_dances | 62 | 5 |
| taking_part_in_clubs | 42 | 7 |
| going_to_fast_foods | 73 | 4 |
| going_to_bars | 71 | 5 |
| smoking | 64 | 4 |
| alcohol_use | 61 | 3 |
| excessive_drunkenness | 69 | 3 |
| sex_first_time | 54 | 52 |

**legend:** Number of discordant twin pairs ranged 42-94. Number of twin pairs available: 257. NA: missing values.

**Table S2:** Associations between within-pair proximity scores and discordance for multiple lifestyle variables.

| | | | | | P-values | | |
|---|---|---|---|---|---|---|---|
| Domain | variable | estimate | se | t-value | nominal | FDR | Bonferroni |
| EAA | vocational_degree | -0.02 | 0.04 | -0.7 | 5.0E-01 | 6.4E-01 | 1.0E+00 |
| EAA | watching_videos | -0.02 | 0.03 | -0.8 | 4.1E-01 | 6.1E-01 | 1.0E+00 |
| EAA | gaming | -0.01 | 0.03 | -0.3 | 7.5E-01 | 8.1E-01 | 1.0E+00 |
| EAA | instrument_playing | -0.05 | 0.03 | -1.3 | 1.8E-01 | 6.1E-01 | 1.0E+00 |
| EAA | reading | 0.00 | 0.03 | 0.1 | 9.4E-01 | 9.4E-01 | 1.0E+00 |
| EAA | going_out | -0.03 | 0.03 | -0.9 | 3.6E-01 | 6.1E-01 | 1.0E+00 |
| EAA | going_to_dances | -0.02 | 0.03 | -0.5 | 6.5E-01 | 7.6E-01 | 1.0E+00 |
| EAA | taking_part_in_clubs | 0.04 | 0.04 | 1.0 | 3.0E-01 | 6.1E-01 | 1.0E+00 |
| EAA | going_to_fast_foods | 0.04 | 0.03 | 1.3 | 1.9E-01 | 6.1E-01 | 1.0E+00 |
| EAA | going_to_bars | -0.05 | 0.03 | -1.4 | 1.6E-01 | 6.1E-01 | 1.0E+00 |
| EAA | smoking | -0.10 | 0.03 | -3.1 | 1.9E-03 | 2.6E-02 | 2.6E-02 |
| EAA | alcohol_use | 0.04 | 0.03 | 1.2 | 2.5E-01 | 6.1E-01 | 1.0E+00 |
| EAA | excessive_drunkenness | 0.03 | 0.03 | 1.0 | 3.2E-01 | 6.1E-01 | 1.0E+00 |
| EAA | sex_first_time | 0.03 | 0.03 | 0.8 | 4.4E-01 | 6.1E-01 | 1.0E+00 |
| Proteome | vocational_degree | -0.02 | 0.02 | -1.0 | 3.1E-01 | 5.3E-01 | 1.0E+00 |
| Proteome | watching_videos | 0.00 | 0.01 | 0.2 | 8.6E-01 | 9.3E-01 | 1.0E+00 |
| Proteome | gaming | -0.03 | 0.01 | -2.0 | 4.7E-02 | 3.3E-01 | 6.6E-01 |
| Proteome | instrument_playing | -0.02 | 0.02 | -1.1 | 2.9E-01 | 5.3E-01 | 1.0E+00 |
| Proteome | reading | -0.02 | 0.01 | -1.5 | 1.3E-01 | 5.1E-01 | 1.0E+00 |
| Proteome | going_out | 0.01 | 0.01 | 0.7 | 5.0E-01 | 6.3E-01 | 1.0E+00 |
| Proteome | going_to_dances | 0.02 | 0.02 | 1.5 | 1.5E-01 | 5.1E-01 | 1.0E+00 |
| Proteome | taking_part_in_clubs | 0.00 | 0.02 | 0.0 | 9.7E-01 | 9.7E-01 | 1.0E+00 |

| | | | | | | | |
|---|---|---|---|---|---|---|---|
| Proteome | going_to_fast_foods | -0.01 | 0.01 | -0.9 | 3.6E-01 | 5.5E-01 | 1.0E+00 |
| Proteome | going_to_bars | -0.01 | 0.02 | -0.4 | 7.3E-01 | 8.5E-01 | 1.0E+00 |
| Proteome | smoking | -0.02 | 0.02 | -1.3 | 2.0E-01 | 5.3E-01 | 1.0E+00 |
| Proteome | alcohol_use | -0.01 | 0.02 | -0.9 | 3.9E-01 | 5.5E-01 | 1.0E+00 |
| Proteome | excessive_drunkenness | -0.04 | 0.02 | -2.5 | 1.2E-02 | 1.6E-01 | 1.6E-01 |
| Proteome | sex_first_time | 0.02 | 0.02 | 1.2 | 2.4E-01 | 5.3E-01 | 1.0E+00 |
| Metabolome | vocational_degree | -0.04 | 0.03 | -1.1 | 2.9E-01 | 8.0E-01 | 1.0E+00 |
| Metabolome | watching_videos | -0.02 | 0.03 | -0.8 | 4.2E-01 | 8.0E-01 | 1.0E+00 |
| Metabolome | gaming | -0.05 | 0.03 | -2.0 | 4.6E-02 | 6.4E-01 | 6.4E-01 |
| Metabolome | instrument_playing | 0.01 | 0.03 | 0.4 | 7.1E-01 | 9.3E-01 | 1.0E+00 |
| Metabolome | reading | -0.02 | 0.03 | -0.7 | 4.6E-01 | 8.0E-01 | 1.0E+00 |
| Metabolome | going_out | -0.02 | 0.03 | -0.7 | 5.1E-01 | 8.0E-01 | 1.0E+00 |
| Metabolome | going_to_dances | -0.03 | 0.03 | -0.9 | 3.5E-01 | 8.0E-01 | 1.0E+00 |
| Metabolome | taking_part_in_clubs | -0.03 | 0.03 | -0.8 | 4.0E-01 | 8.0E-01 | 1.0E+00 |
| Metabolome | going_to_fast_foods | 0.00 | 0.03 | -0.1 | 9.3E-01 | 9.3E-01 | 1.0E+00 |
| Metabolome | going_to_bars | 0.00 | 0.03 | 0.1 | 9.3E-01 | 9.3E-01 | 1.0E+00 |
| Metabolome | smoking | 0.02 | 0.03 | 0.7 | 5.2E-01 | 8.0E-01 | 1.0E+00 |
| Metabolome | alcohol_use | 0.01 | 0.03 | 0.2 | 8.6E-01 | 9.3E-01 | 1.0E+00 |
| Metabolome | excessive_drunkenness | -0.04 | 0.03 | -1.3 | 1.9E-01 | 8.0E-01 | 1.0E+00 |
| Metabolome | sex_first_time | 0.01 | 0.03 | 0.2 | 8.7E-01 | 9.3E-01 | 1.0E+00 |
| External Exposome | vocational_degree | -0.10 | 0.04 | -2.3 | 2.2E-02 | 1.0E-01 | 3.1E-01 |
| External Exposome | watching_videos | -0.05 | 0.03 | -1.5 | 1.3E-01 | 3.7E-01 | 1.0E+00 |
| External Exposome | gaming | 0.03 | 0.04 | 0.8 | 4.1E-01 | 6.3E-01 | 1.0E+00 |

| | | | | | | | |
|---|---|---|---|---|---|---|---|
| External Exposome | instrument_playing | 0.02 | 0.04 | 0.6 | 5.8E-01 | 7.9E-01 | 1.0E+00 |
| External Exposome | reading | -0.01 | 0.03 | -0.3 | 7.3E-01 | 7.9E-01 | 1.0E+00 |
| External Exposome | going_out | -0.10 | 0.03 | -2.8 | 5.1E-03 | 3.5E-02 | 7.1E-02 |
| External Exposome | going_to_dances | -0.05 | 0.04 | -1.2 | 2.4E-01 | 5.7E-01 | 1.0E+00 |
| External Exposome | taking_part_in_clubs | 0.02 | 0.05 | 0.4 | 7.2E-01 | 7.9E-01 | 1.0E+00 |
| External Exposome | going_to_fast_foods | -0.04 | 0.04 | -1.0 | 3.2E-01 | 6.0E-01 | 1.0E+00 |
| External Exposome | going_to_bars | -0.02 | 0.04 | -0.4 | 6.8E-01 | 7.9E-01 | 1.0E+00 |
| External Exposome | smoking | -0.08 | 0.04 | -2.0 | 4.4E-02 | 1.5E-01 | 6.1E-01 |
| External Exposome | alcohol_use | 0.04 | 0.04 | 0.9 | 3.5E-01 | 6.0E-01 | 1.0E+00 |
| External Exposome | excessive_drunkenness | -0.15 | 0.04 | -4.1 | 6.8E-05 | 9.5E-04 | 9.5E-04 |
| External Exposome | sex_first_time | -0.01 | 0.04 | -0.2 | 8.1E-01 | 8.1E-01 | 1.0E+00 |

**legend:** se: standard error.

**Table S3:** Description of variables in the proteome domain

| UniProt ID | *Coding genes* | Protein descriptions |
|---|---|---|
| A1L4H1 | *SSC5D* | Soluble scavenger receptor cysteine-rich domain-containing protein SSC5D |
| O00187 | *MASP2* | Mannan-binding lectin serine protease 2 |
| O00391 | *QSOX1* | Sulfhydryl oxidase 1 |
| O00533 | *CHL1* | Neural cell adhesion molecule L1-like protein |
| O00592 | *PODXL* | Podocalyxin |
| O00602 | *FCN1* | Ficolin-1 |
| O14786 | *NRP1* | Neuropilin-1 |
| O14791 | *APOL1* | Apolipoprotein L1 |
| O15143 | *ARPC1B* | Actin-related protein 2/3 complex subunit 1B |
| O15204 | *ADAMDEC1* | ADAM DEC1 |
| O43493 | *TGOLN2* | Trans-Golgi network integral membrane protein 2 |
| O43505 | *B4GAT1* | Beta-1,4-glucuronyltransferase 1 |
| O43852 | *CALU* | Calumenin |
| O43866 | *CD5L* | CD5 antigen-like |
| O75144 | *ICOSLG* | ICOS ligand |
| O75594 | *PGLYRP1* | Peptidoglycan recognition protein 1 |
| O75636 | *FCN3* | Ficolin-3 |
| O75882 | *ATRN* | Attractin |
| O94919 | *ENDOD1* | Endonuclease domain-containing 1 protein |
| O94985 | *CLSTN1* | Calsyntenin-1 |
| O95445 | *APOM* | Apolipoprotein M |
| O95479 | *H6PD* | GDH/6PGL endoplasmic bifunctional protein |
| O95497 | *VNN1* | Pantetheinase |
| O95998 | *IL18BP* | Interleukin-18-binding protein |
| P00338 | *LDHA* | L-lactate dehydrogenase A chain |
| P00441 | *SOD1* | Superoxide dismutase [Cu-Zn] |
| P00450 | *CP* | Ceruloplasmin |
| P00488 | *F13A1* | Coagulation factor XIII A chain |
| P00533 | *EGFR* | Epidermal growth factor receptor |
| P00734 | *F2* | Prothrombin |

| | | |
|---|---|---|
| P00736 | *C1R* | Complement C1r subcomponent |
| P00740 | *F9* | Coagulation factor IX |
| P00742 | *F10* | Coagulation factor X |
| P00746 | *CFD* | Complement factor D |
| P00747 | *PLG* | Plasminogen |
| P00748 | *F12* | Coagulation factor XII |
| P00751 | *CFB* | Complement factor B |
| P00915 | *CA1* | Carbonic anhydrase 1 |
| P00918 | *CA2* | Carbonic anhydrase 2 |
| P01008 | *SERPINC1* | Antithrombin-III |
| P01011 | *SERPINA3* | Alpha-1-antichymotrypsin |
| P01019 | *AGT* | Angiotensinogen |
| P01024 | *C3* | Complement C3 |
| P01031 | *C5* | Complement C5 |
| P01033 | *TIMP1* | Metalloproteinase inhibitor 1 |
| P01034 | *CST3* | Cystatin-C |
| P01042 | *KNG1* | Kininogen-1 |
| P01137 | *TGFB1* | Transforming growth factor beta-1 proprotein |
| P01344 | *IGF2* | Insulin-like growth factor II |
| P02042 | *HBD* | Hemoglobin subunit delta |
| P02144 | *MB* | Myoglobin |
| P02452 | *COL1A1* | Collagen alpha-1(I) chain |
| P02533 | *KRT14* | Keratin, type I cytoskeletal 14 |
| P02649 | *APOE* | Apolipoprotein E |
| P02652 | *APOA2* | Apolipoprotein A-II |
| P02654 | *APOC1* | Apolipoprotein C-I |
| P02655 | *APOC2* | Apolipoprotein C-II |
| P02656 | *APOC3* | Apolipoprotein C-III |
| P02741 | *CRP* | C-reactive protein |
| P02743 | *APCS* | Serum amyloid P-component |
| P02745 | *C1QA* | Complement C1q subcomponent subunit A |
| P02746 | *C1QB* | Complement C1q subcomponent subunit B |
| P02747 | *C1QC* | Complement C1q subcomponent subunit C |
| P02748 | *C9* | Complement component C9 |
| P02749 | *APOH* | Beta-2-glycoprotein 1 |
| P02750 | *LRG1* | Leucine-rich alpha-2-glycoprotein |
| P02751 | *FN1* | Fibronectin |

| | | |
|---|---|---|
| P02753 | *RBP4* | Retinol-binding protein 4 |
| P02760 | *AMBP* | Protein AMBP |
| P02765 | *AHSG* | Alpha-2-HS-glycoprotein |
| P02766 | *TTR* | Transthyretin |
| P02774 | *GC* | Vitamin D-binding protein |
| P02775 | *PPBP* | Platelet basic protein |
| P02776 | *PF4* | Platelet factor 4 |
| P02790 | *HPX* | Hemopexin |
| P03950 | *ANG* | Angiogenin |
| P03951 | *F11* | Coagulation factor XI |
| P03952 | *KLKB1* | Plasma kallikrein |
| P03973 | *SLPI* | Antileukoproteinase |
| P04003 | *C4BPA* | C4b-binding protein alpha chain |
| P04004 | *VTN* | Vitronectin |
| P04040 | *CAT* | Catalase |
| P04066 | *FUCA1* | Tissue alpha-L-fucosidase |
| P04070 | *PROC* | Vitamin K-dependent protein C |
| P04075 | *ALDOA* | Fructose-bisphosphate aldolase A |
| P04114 | *APOB* | Apolipoprotein B-100 |
| P04180 | *LCAT* | Phosphatidylcholine-sterol acyltransferase |
| P04196 | *HRG* | Histidine-rich glycoprotein |
| P04217 | *A1BG* | Alpha-1B-glycoprotein |
| P04259 | *KRT6B* | Keratin, type II cytoskeletal 6B |
| P04264 | *KRT1* | Keratin, type II cytoskeletal 1 |
| P04275 | *VWF* | von Willebrand factor |
| P04278 | *SHBG* | Sex hormone-binding globulin |
| P04406 | *GAPDH* | Glyceraldehyde-3-phosphate dehydrogenase |
| P04439 | *HLA-A* | HLA class I histocompatibility antigen, A alpha chain |
| P04746 | *AMY2A* | Pancreatic alpha-amylase |
| P05019 | *IGF1* | Insulin-like growth factor I |
| P05062 | *ALDOB* | Fructose-bisphosphate aldolase B |
| P05067 | *APP* | Amyloid-beta precursor protein |
| P05090 | *APOD* | Apolipoprotein D |
| P05106 | *ITGB3* | Integrin beta-3 |
| P05109 | *S100A8* | Protein S100-A8 |
| P05121 | *SERPINE1* | Plasminogen activator inhibitor 1 |
| P05154 | *SERPINA5* | Plasma serine protease inhibitor |

| | | |
|---|---|---|
| P05155 | *SERPING1* | Plasma protease C1 inhibitor |
| P05156 | *CFI* | Complement factor I |
| P05160 | *F13B* | Coagulation factor XIII B chain |
| P05164 | *MPO* | Myeloperoxidase |
| P05362 | *ICAM1* | Intercellular adhesion molecule 1 |
| P05451 | *REG1A* | Lithostathine-1-alpha |
| P05452 | *CLEC3B* | Tetranectin |
| P05543 | *SERPINA7* | Thyroxine-binding globulin |
| P05546 | *SERPIND1* | Heparin cofactor 2 |
| P05556 | *ITGB1* | Integrin beta-1 |
| P06276 | *BCHE* | Cholinesterase |
| P06396 | *GSN* | Gelsolin |
| P06681 | *C2* | Complement C2 |
| P06702 | *S100A9* | Protein S100-A9 |
| P06727 | *APOA4* | Apolipoprotein A-IV |
| P06732 | *CKM* | Creatine kinase M-type |
| P06733 | *ENO1* | Alpha-enolase |
| P07195 | *LDHB* | L-lactate dehydrogenase B chain |
| P07225 | *PROS1* | Vitamin K-dependent protein S |
| P07237 | *P4HB* | Protein disulfide-isomerase |
| P07307 | *ASGR2* | Asialoglycoprotein receptor 2 |
| P07333 | *CSF1R* | Macrophage colony-stimulating factor 1 receptor |
| P07339 | *CTSD* | Cathepsin D |
| P07357 | *C8A* | Complement component C8 alpha chain |
| P07358 | *C8B* | Complement component C8 beta chain |
| P07359 | *GP1BA* | Platelet glycoprotein Ib alpha chain |
| P07360 | *C8G* | Complement component C8 gamma chain |
| P07437 | *TUBB* | Tubulin beta chain |
| P07602 | *PSAP* | Prosaposin |
| P07737 | *PFN1* | Profilin-1 |
| P07911 | *UMOD* | Uromodulin |
| P07942 | *LAMB1* | Laminin subunit beta-1 |
| P07951 | *TPM2* | Tropomyosin beta chain |
| P07996 | *THBS1* | Thrombospondin-1 |
| P07998 | *RNASE1* | Ribonuclease pancreatic |
| P08123 | *COL1A2* | Collagen alpha-2(I) chain |
| P08185 | *SERPINA6* | Corticosteroid-binding globulin |

| | | |
|---|---|---|
| P08195 | *SLC3A2* | 4F2 cell-surface antigen heavy chain |
| P08238 | *HSP90AB1* | Heat shock protein HSP 90-beta |
| P08253 | *MMP2* | 72 kDa type IV collagenase |
| P08294 | *SOD3* | Extracellular superoxide dismutase [Cu-Zn] |
| P08311 | *CTSG* | Cathepsin G |
| P08493 | *MGP* | Matrix Gla protein |
| P08514 | *ITGA2B* | Integrin alpha-IIb |
| P08519 | *LPA* | Apolipoprotein(a) |
| P08571 | *CD14* | Monocyte differentiation antigen CD14 |
| P08603 | *CFH* | Complement factor H |
| P08697 | *SERPINF2* | Alpha-2-antiplasmin |
| P08709 | *F7* | Coagulation factor VII |
| P08779 | *KRT16* | Keratin, type I cytoskeletal 16 |
| P08887 | *IL6R* | Interleukin-6 receptor subunit alpha |
| P09172 | *DBH* | Dopamine beta-hydroxylase |
| P09486 | *SPARC* | SPARC |
| P09871 | *C1S* | Complement C1s subcomponent |
| P09960 | *LTA4H* | Leukotriene A-4 hydrolase |
| P0C0L4 | *C4A* | Complement C4-A |
| P0C0L5 | *C4B* | Complement C4-B |
| P0DJI8 | *SAA1* | Serum amyloid A-1 protein |
| P0DJI9 | *SAA2* | Serum amyloid A-2 protein |
| P10124 | *SRGN* | Serglycin |
| P10153 | *RNASE2* | Non-secretory ribonuclease |
| P10586 | *PTPRF* | Receptor-type tyrosine-protein phosphatase F |
| P10643 | *C7* | Complement component C7 |
| P10646 | *TFPI* | Tissue factor pathway inhibitor |
| P10720 | *PF4V1* | Platelet factor 4 variant |
| P10721 | *KIT* | Mast/stem cell growth factor receptor Kit |
| P10909 | *CLU* | Clusterin |
| P11021 | *HSPA5* | Endoplasmic reticulum chaperone BiP |
| P11047 | *LAMC1* | Laminin subunit gamma-1 |
| P11142 | *HSPA8* | Heat shock cognate 71 kDa protein |
| P11226 | *MBL2* | Mannose-binding protein C |
| P11279 | *LAMP1* | Lysosome-associated membrane glycoprotein 1 |
| P11362 | *FGFR1* | Fibroblast growth factor receptor 1 |
| P11597 | *CETP* | Cholesteryl ester transfer protein |

| | | |
|---|---|---|
| P11717 | *IGF2R* | Cation-independent mannose-6-phosphate receptor |
| P12109 | *COL6A1* | Collagen alpha-1(VI) chain |
| P12111 | *COL6A3* | Collagen alpha-3(VI) chain |
| P12259 | *F5* | Coagulation factor V |
| P12821 | *ACE* | Angiotensin-converting enzyme |
| P12830 | *CDH1* | Cadherin-1 |
| P12955 | *PEPD* | Xaa-Pro dipeptidase |
| P13473 | *LAMP2* | Lysosome-associated membrane glycoprotein 2 |
| P13591 | *NCAM1* | Neural cell adhesion molecule 1 |
| P13598 | *ICAM2* | Intercellular adhesion molecule 2 |
| P13645 | *KRT10* | Keratin, type I cytoskeletal 10 |
| P13646 | *KRT13* | Keratin, type I cytoskeletal 13 |
| P13671 | *C6* | Complement component C6 |
| P13727 | *PRG2* | Bone marrow proteoglycan |
| P13796 | *LCP1* | Plastin-2 |
| P14151 | *SELL* | L-selectin |
| P14209 | *CD99* | CD99 antigen |
| P14314 | *PRKCSH* | Glucosidase 2 subunit beta |
| P14543 | *NID1* | Nidogen-1 |
| P14618 | *PKM* | Pyruvate kinase PKM |
| P14625 | *HSP90B1* | Endoplasmin |
| P14780 | *MMP9* | Matrix metalloproteinase-9 |
| P15144 | *ANPEP* | Aminopeptidase N |
| P15151 | *PVR* | Poliovirus receptor |
| P15169 | *CPN1* | Carboxypeptidase N catalytic chain |
| P15291 | *B4GALT1* | Beta-1,4-galactosyltransferase 1 |
| NA | *NA* | NA |
| P16035 | *TIMP2* | Metalloproteinase inhibitor 2 |
| P16070 | *CD44* | CD44 antigen |
| P16109 | *SELP* | P-selectin |
| P16112 | *ACAN* | Aggrecan core protein |
| P16930 | *FAH* | Fumarylacetoacetase |
| P17813 | *ENG* | Endoglin |
| P17936 | *IGFBP3* | Insulin-like growth factor-binding protein 3 |
| P18065 | *IGFBP2* | Insulin-like growth factor-binding protein 2 |
| P18206 | *VCL* | Vinculin |
| P18428 | *LBP* | Lipopolysaccharide-binding protein |

| | | |
|---|---|---|
| P19021 | *PAM* | Peptidyl-glycine alpha-amidating monooxygenase |
| P19022 | *CDH2* | Cadherin-2 |
| P19320 | *VCAM1* | Vascular cell adhesion protein 1 |
| P19823 | *ITIH2* | Inter-alpha-trypsin inhibitor heavy chain H2 |
| P19827 | *ITIH1* | Inter-alpha-trypsin inhibitor heavy chain H1 |
| P20023 | *CR2* | Complement receptor type 2 |
| P20742 | *PZP* | Pregnancy zone protein |
| P20851 | *C4BPB* | C4b-binding protein beta chain |
| P20930 | *FLG* | Filaggrin |
| P21333 | *FLNA* | Filamin-A |
| P21926 | *CD9* | CD9 antigen |
| P22105 | *TNXB* | Tenascin-X |
| P22352 | *GPX3* | Glutathione peroxidase 3 |
| P22692 | *IGFBP4* | Insulin-like growth factor-binding protein 4 |
| P22792 | *CPN2* | Carboxypeptidase N subunit 2 |
| P22891 | *PROZ* | Vitamin K-dependent protein Z |
| P22897 | *MRC1* | Macrophage mannose receptor 1 |
| P23142 | *FBLN1* | Fibulin-1 |
| P23470 | *PTPRG* | Receptor-type tyrosine-protein phosphatase gamma |
| P23528 | *CFL1* | Cofilin-1 |
| P24043 | *LAMA2* | Laminin subunit alpha-2 |
| P24592 | *IGFBP6* | Insulin-like growth factor-binding protein 6 |
| P24593 | *IGFBP5* | Insulin-like growth factor-binding protein 5 |
| P24821 | *TNC* | Tenascin |
| P25311 | *AZGP1* | Zinc-alpha-2-glycoprotein |
| P25774 | *CTSS* | Cathepsin S |
| P26038 | *MSN* | Moesin |
| P26927 | *MST1* | Hepatocyte growth factor-like protein |
| P27105 | *STOM* | Stomatin |
| P27169 | *PON1* | Serum paraoxonase/arylesterase 1 |
| P27487 | *DPP4* | Dipeptidyl peptidase 4 |
| P27797 | *CALR* | Calreticulin |
| P27918 | *CFP* | Properdin |
| P28799 | *GRN* | Progranulin |
| P29279 | *CCN2* | CCN family member 2 |
| P29401 | *TKT* | Transketolase |
| P29622 | *SERPINA4* | Kallistatin |

| | | |
|---|---|---|
| P30043 | *BLVRB* | Flavin reductase (NADPH) |
| P30101 | *PDIA3* | Protein disulfide-isomerase A3 |
| P32119 | *PRDX2* | Peroxiredoxin-2 |
| P32942 | *ICAM3* | Intercellular adhesion molecule 3 |
| P33151 | *CDH5* | Cadherin-5 |
| P33908 | *MAN1A1* | Mannosyl-oligosaccharide 1,2-alpha-mannosidase IA |
| P34096 | *RNASE4* | Ribonuclease 4 |
| P35030 | *PRSS3* | Trypsin-3 |
| P35443 | *THBS4* | Thrombospondin-4 |
| P35527 | *KRT9* | Keratin, type I cytoskeletal 9 |
| P35542 | *SAA4* | Serum amyloid A-4 protein |
| P35555 | *FBN1* | Fibrillin-1 |
| P35858 | *IGFALS* | Insulin-like growth factor-binding protein complex acid labile subunit |
| P35908 | *KRT2* | Keratin, type II cytoskeletal 2 epidermal |
| P35916 | *FLT4* | Vascular endothelial growth factor receptor 3 |
| P36955 | *SERPINF1* | Pigment epithelium-derived factor |
| P36980 | *CFHR2* | Complement factor H-related protein 2 |
| P39060 | *COL18A1* | Collagen alpha-1(XVIII) chain |
| P40189 | *IL6ST* | Interleukin-6 receptor subunit beta |
| P40197 | *GP5* | Platelet glycoprotein V |
| P41222 | *PTGDS* | Prostaglandin-H2 D-isomerase |
| P43121 | *MCAM* | Cell surface glycoprotein MUC18 |
| P43251 | *BTD* | Biotinidase |
| P43652 | *AFM* | Afamin |
| P48594 | *SERPINB4* | Serpin B4 |
| P48668 | *KRT6C* | Keratin, type II cytoskeletal 6C |
| P48740 | *MASP1* | Mannan-binding lectin serine protease 1 |
| P49747 | *COMP* | Cartilage oligomeric matrix protein |
| P49908 | *SELENOP* | Selenoprotein P |
| P49913 | *CAMP* | Cathelicidin antimicrobial peptide |
| P50395 | *GDI2* | Rab GDP dissociation inhibitor beta |
| P51884 | *LUM* | Lumican |
| P53634 | *CTSC* | Dipeptidyl peptidase 1 |
| P54289 | *CACNA2D1* | Voltage-dependent calcium channel subunit alpha-2/delta-1 |
| P54764 | *EPHA4* | Ephrin type-A receptor 4 |
| P54802 | *NAGLU* | Alpha-N-acetylglucosaminidase |

| | | |
|---|---|---|
| P55056 | *APOC4* | Apolipoprotein C-IV |
| P55058 | *PLTP* | Phospholipid transfer protein |
| P55103 | *INHBC* | Inhibin beta C chain |
| P55290 | *CDH13* | Cadherin-13 |
| P59666 | *DEFA3* | Neutrophil defensin 3 |
| P60174 | *TPI1* | Triosephosphate isomerase |
| P60709 | *ACTB* | Actin, cytoplasmic 1 |
| P61224 | *RAP1B* | Ras-related protein Rap-1b |
| P61626 | *LYZ* | Lysozyme C |
| P61769 | *B2M* | Beta-2-microglobulin |
| P62328 | *TMSB4X* | Thymosin beta-4 |
| P62913 | *RPL11* | 60S ribosomal protein L11 |
| P62937 | *PPIA* | Peptidyl-prolyl cis-trans isomerase A |
| P63104 | *YWHAZ* | 14-3-3 protein zeta/delta |
| P63261 | *ACTG1* | Actin, cytoplasmic 2 |
| NA | *NA* | NA |
| P68363 | *TUBA1B* | Tubulin alpha-1B chain |
| P68871 | *HBB* | Hemoglobin subunit beta |
| P69892 | *HBG2* | Hemoglobin subunit gamma-2 |
| P69905 | *HBA1* | Hemoglobin subunit alpha |
| P78417 | *GSTO1* | Glutathione S-transferase omega-1 |
| P78509 | *RELN* | Reelin |
| P80108 | *GPLD1* | Phosphatidylinositol-glycan-specific phospholipase D |
| P80723 | *BASP1* | Brain acid soluble protein 1 |
| P98160 | *HSPG2* | Basement membrane-specific heparan sulfate proteoglycan core protein |
| Q00610 | *CLTC* | Clathrin heavy chain 1 |
| Q01459 | *CTBS* | Di-N-acetylchitobiase |
| Q01469 | *FABP5* | Fatty acid-binding protein 5 |
| Q02985 | *CFHR3* | Complement factor H-related protein 3 |
| Q03591 | *CFHR1* | Complement factor H-related protein 1 |
| Q04695 | *KRT17* | Keratin, type I cytoskeletal 17 |
| Q04721 | *NOTCH2* | Neurogenic locus notch homolog protein 2 |
| Q04756 | *HGFAC* | Hepatocyte growth factor activator |
| Q06033 | *ITIH3* | Inter-alpha-trypsin inhibitor heavy chain H3 |
| Q06830 | *PRDX1* | Peroxiredoxin-1 |
| Q07954 | *LRP1* | Prolow-density lipoprotein receptor-related protein 1 |

| | | |
|---|---|---|
| Q08380 | *LGALS3BP* | Galectin-3-binding protein |
| Q10588 | *BST1* | ADP-ribosyl cyclase/cyclic ADP-ribose hydrolase 2 |
| Q12805 | *EFEMP1* | EGF-containing fibulin-like extracellular matrix protein 1 |
| Q12841 | *FSTL1* | Follistatin-related protein 1 |
| Q12860 | *CNTN1* | Contactin-1 |
| Q12884 | *FAP* | Prolyl endopeptidase FAP |
| Q12907 | *LMAN2* | Vesicular integral-membrane protein VIP36 |
| Q12913 | *PTPRJ* | Receptor-type tyrosine-protein phosphatase eta |
| Q13093 | *PLA2G7* | Platelet-activating factor acetylhydrolase |
| Q13103 | *SPP2* | Secreted phosphoprotein 24 |
| Q13201 | *MMRN1* | Multimerin-1 |
| Q13332 | *PTPRS* | Receptor-type tyrosine-protein phosphatase S |
| Q13449 | *LSAMP* | Limbic system-associated membrane protein |
| Q13740 | *ALCAM* | CD166 antigen |
| Q13790 | *APOF* | Apolipoprotein F |
| Q13822 | *ENPP2* | Ectonucleotide pyrophosphatase/phosphodiesterase family member 2 |
| Q14112 | *NID2* | Nidogen-2 |
| Q14126 | *DSG2* | Desmoglein-2 |
| Q14314 | *FGL2* | Fibroleukin |
| Q14515 | *SPARCL1* | SPARC-like protein 1 |
| Q14520 | *HABP2* | Hyaluronan-binding protein 2 |
| Q14624 | *ITIH4* | Inter-alpha-trypsin inhibitor heavy chain H4 |
| Q14766 | *LTBP1* | Latent-transforming growth factor beta-binding protein 1 |
| Q14956 | *GPNMB* | Transmembrane glycoprotein NMB |
| Q15063 | *POSTN* | Periostin |
| Q15113 | *PCOLCE* | Procollagen C-endopeptidase enhancer 1 |
| Q15166 | *PON3* | Serum paraoxonase/lactonase 3 |
| Q15485 | *FCN2* | Ficolin-2 |
| Q15582 | *TGFBI* | Transforming growth factor-beta-induced protein ig-h3 |
| Q15828 | *CST6* | Cystatin-M |
| Q15848 | *ADIPOQ* | Adiponectin |
| Q16270 | *IGFBP7* | Insulin-like growth factor-binding protein 7 |
| Q16610 | *ECM1* | Extracellular matrix protein 1 |
| Q16706 | *MAN2A1* | Alpha-mannosidase 2 |
| Q16853 | *AOC3* | Membrane primary amine oxidase |

| | | |
|---|---|---|
| Q4LDE5 | *SVEP1* | Sushi, von Willebrand factor type A, EGF and pentraxin domain-containing protein 1 |
| Q562R1 | *ACTBL2* | Beta-actin-like protein 2 |
| Q5D862 | *FLG2* | Filaggrin-2 |
| Q6EMK4 | *VASN* | Vasorin |
| Q6UWP8 | *SBSN* | Suprabasin |
| Q6UX71 | *PLXDC2* | Plexin domain-containing protein 2 |
| Q6UXB8 | *PI16* | Peptidase inhibitor 16 |
| Q6UY14 | *ADAMTSL4* | ADAMTS-like protein 4 |
| Q6YHK3 | *CD109* | CD109 antigen |
| Q76LX8 | *ADAMTS13* | A disintegrin and metalloproteinase with thrombospondin motifs 13 |
| Q7Z7G0 | *ABI3BP* | Target of Nesh-SH3 |
| Q7Z7M0 | *MEGF8* | Multiple epidermal growth factor-like domains protein 8 |
| Q86SQ4 | *ADGRG6* | Adhesion G-protein coupled receptor G6 |
| Q86TH1 | *ADAMTSL2* | ADAMTS-like protein 2 |
| Q86U17 | *SERPINA11* | Serpin A11 |
| Q86UD1 | *OAF* | Out at first protein homolog |
| Q86UX7 | *FERMT3* | Fermitin family homolog 3 |
| Q86VB7 | *CD163* | Scavenger receptor cysteine-rich type 1 protein M130 |
| Q86YW5 | *TREML1* | Trem-like transcript 1 protein |
| Q86YZ3 | *HRNR* | Hornerin |
| Q8IUL8 | *CILP2* | Cartilage intermediate layer protein 2 |
| Q8IZF2 | *ADGRF5* | Adhesion G protein-coupled receptor F5 |
| Q8N1F8 | *STK11IP* | Serine/threonine-protein kinase 11-interacting protein |
| Q8N3V7 | *SYNPO* | Synaptopodin |
| Q8NBP7 | *PCSK9* | Proprotein convertase subtilisin/kexin type 9 |
| Q8NDA2 | *HMCN2* | Hemicentin-2 |
| Q8WZ75 | *ROBO4* | Roundabout homolog 4 |
| Q92496 | *CFHR4* | Complement factor H-related protein 4 |
| Q92820 | *GGH* | Gamma-glutamyl hydrolase |
| Q92954 | *PRG4* | Proteoglycan 4 |
| Q96EE4 | *CCDC126* | Coiled-coil domain-containing protein 126 |
| Q96IY4 | *CPB2* | Carboxypeptidase B2 |
| Q96KG7 | *MEGF10* | Multiple epidermal growth factor-like domains protein 10 |
| Q96KN2 | *CNDP1* | Beta-Ala-His dipeptidase |
| Q96NZ9 | *PRAP1* | Proline-rich acidic protein 1 |

| | | |
|---|---|---|
| Q96PD5 | *PGLYRP2* | N-acetylmuramoyl-L-alanine amidase |
| Q96S96 | *PEBP4* | Phosphatidylethanolamine-binding protein 4 |
| Q99784 | *OLFM1* | Noelin |
| Q99969 | *RARRES2* | Retinoic acid receptor responder protein 2 |
| Q99983 | *OMD* | Osteomodulin |
| Q9BTY2 | *FUCA2* | Plasma alpha-L-fucosidase |
| Q9BUN1 | *MENT* | Protein MENT |
| Q9BXJ4 | *C1QTNF3* | Complement C1q tumor necrosis factor-related protein 3 |
| Q9BXR6 | *CFHR5* | Complement factor H-related protein 5 |
| Q9BY67 | *CADM1* | Cell adhesion molecule 1 |
| Q9H1U4 | *MEGF9* | Multiple epidermal growth factor-like domains protein 9 |
| Q9H299 | *SH3BGRL3* | SH3 domain-binding glutamic acid-rich-like protein 3 |
| Q9H4A9 | *DPEP2* | Dipeptidase 2 |
| Q9H4B7 | *TUBB1* | Tubulin beta-1 chain |
| Q9H4G4 | *GLIPR2* | Golgi-associated plant pathogenesis-related protein 1 |
| Q9H8L6 | *MMRN2* | Multimerin-2 |
| Q9HDC9 | *APMAP* | Adipocyte plasma membrane-associated protein |
| Q9NPH3 | *IL1RAP* | Interleukin-1 receptor accessory protein |
| Q9NPR2 | *SEMA4B* | Semaphorin-4B |
| Q9NPY3 | *CD93* | Complement component C1q receptor |
| Q9NQ79 | *CRTAC1* | Cartilage acidic protein 1 |
| Q9NTU7 | *CBLN4* | Cerebellin-4 |
| Q9NY15 | *STAB1* | Stabilin-1 |
| Q9NY97 | *B3GNT2* | N-acetyllactosaminide beta-1,3-N-acetylglucosaminyltransferase 2 |
| Q9NZK5 | *ADA2* | Adenosine deaminase 2 |
| Q9NZP8 | *C1RL* | Complement C1r subcomponent-like protein |
| Q9P232 | *CNTN3* | Contactin-3 |
| Q9UBR2 | *CTSZ* | Cathepsin Z |
| Q9UBX1 | *CTSF* | Cathepsin F |
| Q9UEW3 | *MARCO* | Macrophage receptor MARCO |
| Q9UGM5 | *FETUB* | Fetuin-B |
| Q9UHG3 | *PCYOX1* | Prenylcysteine oxidase 1 |
| Q9UJJ9 | *GNPTG* | N-acetylglucosamine-1-phosphotransferase subunit gamma |
| Q9UK55 | *SERPINA10* | Protein Z-dependent protease inhibitor |
| Q9ULI3 | *HEG1* | Protein HEG homolog 1 |
| Q9UNN8 | *PROCR* | Endothelial protein C receptor |

| | | |
|---|---|---|
| Q9UNW1 | *MINPP1* | Multiple inositol polyphosphate phosphatase 1 |
| Q9Y251 | *HPSE* | Heparanase |
| Q9Y490 | *TLN1* | Talin-1 |
| Q9Y4L1 | *HYOU1* | Hypoxia up-regulated protein 1 |
| Q9Y5C1 | *ANGPTL3* | Angiopoietin-related protein 3 |
| Q9Y5Y7 | *LYVE1* | Lymphatic vessel endothelial hyaluronic acid receptor 1 |
| Q9Y646 | *CPQ* | Carboxypeptidase Q |
| Q9Y6R7 | *FCGBP* | IgGFc-binding protein |
| Q9Y6Z7 | *COLEC10* | Collectin-10 |

**Table S4:** Description of variables in the metabolome domain

| Variable name | Metabolite |
|---|---|
| alb | Albumin |
| xxl_vldl_pl | Phospholipids in chylomicrons and extremely large VLDL |
| xxl_vldl_l | Total lipids in chylomicrons and extremely large VLDL |
| xxl_vldl_p | Concentration of chylomicrons and extremely large VLDL particles |
| xl_vldl_pl | Phospholipids in very large VLDL |
| xl_vldl_tg | Triglycerides in very large VLDL |
| xl_vldl_l | Total lipids in very large VLDL |
| xl_vldl_p | Concentration of very large VLDL particles |
| l_vldl_c | Cholesterol in large VLDL |
| l_vldl_fc | Free cholesterol in large VLDL |
| l_vldl_pl | Phospholipids in large VLDL |
| l_vldl_tg | Triglycerides in large VLDL |
| l_vldl_ce | Cholesteryl esters in large VLDL |
| l_vldl_l | Total lipids in large VLDL |
| l_vldl_p | Concentration of large VLDL particles |
| m_vldl_c | Cholesterol in medium VLDL |
| m_vldl_fc | Free cholesterol in medium VLDL |
| m_vldl_pl | Phospholipids in medium VLDL |
| m_vldl_tg | Triglycerides in medium VLDL |
| m_vldl_ce | Cholesteryl esters in medium VLDL |
| m_vldl_l | Total lipids in medium VLDL |
| m_vldl_p | Concentration of medium VLDL particles |
| s_vldl_c | Cholesterol in small VLDL |
| s_vldl_fc | Free cholesterol in small VLDL |
| s_vldl_pl | Phospholipids in small VLDL |
| s_vldl_tg | Triglycerides in small VLDL |
| s_vldl_l | Total lipids in small VLDL |
| s_vldl_p | Concentration of small VLDL particles |
| xs_vldl_pl | Phospholipids in very small VLDL |
| xs_vldl_tg | Triglycerides in very small VLDL |
| xs_vldl_l | Total lipids in very small VLDL |
| xs_vldl_p | Concentration of very small VLDL particles |
| idl_fc | Free cholesterol in IDL |
| idl_pl | Phospholipids in IDL |

| | |
|---|---|
| idl_l | Total lipids in IDL |
| idl_p | Concentration of IDL particles |
| l_ldl_c | Cholesterol in large LDL |
| l_ldl_fc | Free cholesterol in large LDL |
| l_ldl_pl | Phospholipids in large LDL |
| l_ldl_ce | Cholesteryl esters in large LDL |
| l_ldl_l | Total lipids in large LDL |
| l_ldl_p | Concentration of large LDL particles |
| m_ldl_c | Cholesterol in medium LDL |
| m_ldl_pl | Phospholipids in medium LDL |
| m_ldl_ce | Cholesteryl esters in medium LDL |
| m_ldl_l | Total lipids in medium LDL |
| m_ldl_p | Concentration of medium LDL particles |
| s_ldl_c | Cholesterol in small LDL |
| s_ldl_l | Total lipids in small LDL |
| s_ldl_p | Concentration of small LDL particles |
| xl_hdl_c | Cholesterol in very large HDL |
| xl_hdl_fc | Free cholesterol in very large HDL |
| xl_hdl_pl | Phospholipids in very large HDL |
| xl_hdl_tg | Triglycerides in very large HDL |
| xl_hdl_ce | Cholesteryl esters in very large HDL |
| xl_hdl_l | Total lipids in very large HDL |
| xl_hdl_p | Concentration of very large HDL particles |
| l_hdl_c | Cholesterol in large HDL |
| l_hdl_fc | Free cholesterol in large HDL |
| l_hdl_pl | Phospholipids in large HDL |
| l_hdl_ce | Cholesteryl esters in large HDL |
| l_hdl_l | Total lipids in large HDL |
| l_hdl_p | Concentration of large HDL particles |
| m_hdl_c | Cholesterol in medium HDL |
| m_hdl_fc | Free cholesterol in medium HDL |
| m_hdl_pl | Phospholipids in medium HDL |
| m_hdl_ce | Cholesteryl esters in medium HDL |
| m_hdl_l | Total lipids in medium HDL |
| m_hdl_p | Concentration of medium HDL particles |
| s_hdl_tg | Triglycerides in small HDL |
| s_hdl_l | Total lipids in small HDL |

| | |
|---|---|
| s_hdl_p | Concentration of small HDL particles |
| xxl_vldl_tg | Triglycerides in chylomicrons and extremely large VLDL |
| vldl_tg | Triglycerides in VLDL |
| idl_tg | Triglycerides in IDL |
| idl_c | Cholesterol in IDL |
| ldl_c | LDL cholesterol |
| hdl_c | HDL cholesterol |
| serum_tg | Serum total triglycerides |
| serum_c | Serum total cholesterol |
| vldl_d | Average diameter for VLDL particles |
| ldl_d | Average diameter for LDL particles |
| hdl_d | Average diameter for HDL particles |
| vldl_tg_efr | Triglycerides in VLDL EFR |
| idl_c_efr | Cholesterol in IDL EFR |
| ldl_c_efr | LDL cholesterol EFR |
| hdl2_c | HDL cholesterol subfraction 2 |
| apoa1 | Apolipoprotein A1 |
| apob | Apolipoprotein B |
| apobtoapoa1 | Ratio of apolipoprotein B to apolipoprotein A1 |
| hdl3_c | HDL cholesterol subfraction 3 |
| bohbut | 3-Hydroxybutyrate |
| ace | Acetate |
| acace | Acetoacetate |
| ala | Alanine |
| mobch2 | CH2 groups of mobile lipids |
| mobch3 | CH3 groups of mobile lipids |
| cit | Citrate |
| crea | Creatinine |
| mobch | CH groups of mobile lipids |
| glc | Glucose |
| gln | Glutamine |
| glol | Glycerol |
| gp | Glycoprotein acetyls, mainly a1-acid glycoprotein |
| his | Histidine |
| ile | Isoleucine |
| lac | Lactate |
| leu | Leucine |

| | |
|---|---|
| phe | Phenylalanine |
| pyr | Pyruvate |
| tyr | Tyrosine |
| urea | Urea |
| val | Valine |
| ldl_friedewald | LDL Friedewald |
| EstC | Esterified cholesterol |
| FreeC | Free cholesterol |
| FAw3 | Omega-3 fatty acids |
| FAw6 | Omega-6 fatty acids |
| FAw79S | Omega-7 and -9 and saturated fatty acids |
| TotFA | Total fatty acids |
| la | Linoleic acid |
| otPUFA | Other polyunsaturated fatty acids than 18:2 |
| dha | Docosahexaenoic acid |
| mufa | Monounsaturated fatty acids |
| TotPG | Total phosphoglycerides |
| pc | Phosphatidylcholine and other cholines |
| sm | Sphingomyelins |
| FAw3toFA | Ratio of omega-3 fatty acids to total fatty acids |
| FAw6toFA | Ratio of omega-6 fatty acids to total fatty acids |
| FAw79StoFA | Ratio of FAw79S to total fatty acids |
| CH2inFA | CH2 groups in fatty acids |
| TGtoPG | Ratio of triglycerides to phosphoglycerides |
| CH2toDB | Ratio of CH2 groups in fatty acids to phosphoglycerides |
| DBinFA | Double bonds in fatty acids |
| BIStoDB | Ratio of bis-allylic bonds to double bonds in lipids |
| BIStoFA | Ratio of bis-allylic bonds to total fatty acids in lipids |
| FALen | Estimated description of fatty acid chain length, not actual carbon number |
| LAtoFA | Ratio of 18:2 linoleic acid to total fatty acids |
| MUFAtoFA | Ratio of monounsaturated fatty acids to total fatty acids |
| DHAtoFA | Ratio of 22:6 docosahexaenoic acid to total fatty acids |

**Table S5:** Description of variables in the external exposome domain

| Exposure ID | Description |
|---|---|
| age0_2006 | Age structure (10-year group): proportion of people age between 0 and 9 in the total population |
| age10_2006 | Age structure (10-year group): proportion of people age between 10 and 19 in the total population |
| age20_2006 | Age structure (10-year group): proportion of people age between 20 and 29 in the total population |
| age30_2006 | Age structure (10-year group): proportion of people age between 30 and 39 in the total population |
| age40_2006 | Age structure (10-year group): proportion of people age between 40 and 49 in the total population |
| age50_2006 | Age structure (10-year group): proportion of people age between 50 and 59 in the total population |
| age60_2006 | Age structure (10-year group): proportion of people age between 60 and 69 in the total population |
| age70_2006 | Age structure (10-year group): proportion of people age between 70 and 79 in the total population |
| age80_2006 | Age structure (10-year group): proportion of people age between 80 and 89 in the total population |
| age90_2006 | Age structure (10-year group): proportion of people age over 90 in the total population |
| under66_2006 | Age structure (old-age ratio): proportion of people age under 66 in the total population |
| over66_2006 | Age structure (old-age ratio): proportion of people age over 66 in the total population |
| under18_2006 | Age structure (youth ratio): proportion of people age under 18 in the total population |
| over18_2006 | Age structure (youth ratio): proportion of people age over 18 in the total population |
| edu1_2006 | Education level: unknown or no education after primary or lower secondary education of the population aged 16 and above (%) |
| edu2_2006 | Education level: upper secondary education (ISCED 3) of the population aged 16 and above (%) |
| edu3_2006 | Education level: post-secondary non-tertiary education / Short-cycle tertiary education (ISCED 4-5) of the population aged 16 and above (%) |
| edu4_2006 | Education level: bachelor′s/equivalent (ISCED 6) of the population aged 16 and above (%) |
| edu5_2006 | Education level: master′s/equivalent (ISCED 7) of the population aged 16 and above (%) |

| | |
|---|---|
| edu6_2006 | Education level: doctoral/equivalent (ISCED 8) of the population aged 16 and above (%) |
| unemp_2006 | Unemployment rates of the total population (%) |
| unemp2554_2006 | Unemployment rates among people who were between 25 and 54 years old and older (%) |
| unemp55_2006 | Unemployment rates among people who were 55 years old and older (%) |
| unemp1824_2006 | Unemployment rates among people who were between 18 and 24 years old (%) |
| unempm_2006 | Unemployment rates among males (%) |
| unempw_2006 | Unemployment rates among females (%) |
| background1_2006 | Percentage of people, born in Finland, with foreign background in the total population |
| background2_2006 | Percentage of people, born abroad, with foreign background in the total population |
| fcitizen_2006 | Percentage of people with foreign citizenship in the total population |
| evluth_2006 | Percentage of people belonging to Lutheran communities in the total population |
| other_2006 | Percentage of people belonging to other religious communities in the total population |
| type1_2006 | Stage in life/structure of household-dwelling units: single households (%) |
| type2_2006 | Stage in life/structure of household-dwelling units: single parent with underage child/children (%) |
| type3_2006 | Stage in life/structure of household-dwelling units: married/registered/cohabiting couples without underage child/children (%) |
| type4_2006 | Stage in life/structure of household-dwelling units: married/registered/cohabiting couples with underage child/children (%) |
| type5_2006 | Stage in life/structure of household-dwelling units: other households (eg. family and a grandparent (or other single person); several families; several single persons; couple with older child/children; single parent with older child/children) (%) |
| finsa_2006 | Proportion of household-dwelling units with an official language as Finnish and Sami in all households |
| swe_2006 | Proportion of household-dwelling units with an official language as Swedish in all households |
| quartile1_2006 | Proportion of households in first quartile (lowest) income level in the country |
| quartile2_2006 | Proportion of households in second quartile income level in the country |

| | |
|---|---|
| quartile3_2006 | Proportion of households in third quartile income level in the country |
| quartile4_2006 | Proportion of households in forth quartile (highest) income level in the country |
| netmig_2006 | Net migration rate: difference between the number of people entering (migration) an area and people leaving (emigration) an area |
| vote_mun_2004 | Voting turnout of municipal election (%)(municipality-level) |
| crime11_2006 | Crime rate per capita: offences known to the authorities per 1000 population in the area, category: offences against property (municipality-level) |
| crime12_2006 | Crime rate per capita: offences known to the authorities per 1000 population in the area, category: crimes against life and health (municipality-level) |
| crime13_2006 | Crime rate per capita: offences known to the authorities per 1000 population in the area, category: sexual crimes (municipality-level) |
| crime14_2006 | Crime rate per capita: offences known to the authorities per 1000 population in the area, category: crimes against public authority and public peace (municipality-level) |
| crime15_2006 | Crime rate per capita: offences known to the authorities per 1000 population in the area, category: traffic offences (municipality-level) |
| crime16_2006 | Crime rate per capita: offences known to the authorities per 1000 population in the area, category: other offences against the penal code (municipality-level) |
| crime02_2006 | Crime rate per capita: offences known to the authorities per 1000 population in the area, category:other acts (municipality-level) |
| vote_par_2007 | Voting turnout of parliamentary election (%)(municipality-level) |
| built_isa_100_2006 | Percentage (%) of built up area within a buffer area of 100m radius around twins' residence, source: ISA. |
| built_isa_300_2006 | Percentage (%) of built up area within a buffer area of 300m radius around twins' residence, source: ISA. |
| built_isa_500_2006 | Percentage (%) of built up area within a buffer area of 500m radius around twins' residence, source: ISA. |
| dist_green_clc_2006 | Euclidean distance to closer green space (unit: m), source: CLC. |
| size_green_clc_2006 | Area of the closer green space (unit: m2), source: CLC. |
| dist_blue_clc_2006 | Euclidean distance to closer blue space (unit: m), source: CLC. |
| size_blue_clc_ | Area of the closer blue space (unit: m2), source: CLC. |

| | |
|---|---|
| 2006 | |
| treecover_2005_100 | Percentage of area covered by trees within a 100 meter buffer around geocode |
| treecover_2005_300 | Percentage of area covered by trees within a 300 meter buffer around geocode |
| treecover_2005_500 | Percentage of area covered by trees within a 500 meter buffer around geocode |
| msavi_5yrs_all_100_2006 | 5-years moving average (2002 to 2006) of MSAVI within a 100 meter buffer around geocode using all available images during the whole year. |
| msavi_5yrs_all_300_2006 | 5-years moving average (2002 to 2006) of MSAVI within a 300 meter buffer around geocode using all available images during the whole year. |
| msavi_5yrs_all_500_2006 | 5-years moving average (2002 to 2006) of MSAVI within a 500 meter buffer around geocode using all available images during the whole year. |
